# The Economic Costs of Child Maltreatment in UK

**DOI:** 10.1101/2021.07.24.21261071

**Authors:** Gabriella Conti, Elena Pizzo, Stephen Morris, Mariya Melnychuk

**Author notes:** Gabriella Conti: Drayton House, 30 Gordon Street, London WC1H 0AX, United Kingdom.

## Abstract

Child maltreatment is a major public health problem with significant consequences for individual victims and for society. In this paper we quantify for the first time the economic costs of fatal and non-fatal child maltreatment in the UK in relation to several short-, medium- and long-term outcomes ranging from physical and mental health problems, to labour market outcomes and welfare use. We combine novel regression analysis of rich data from the National Child Development Study and the English Longitudinal Study of Ageing with secondary evidence to produce an incidence-based estimate of the lifetime costs of child maltreatment from a societal perspective. The discounted average lifetime incidence cost of non-fatal child maltreatment by a primary caregiver is estimated at £89,390 (95% uncertainty interval £44,896 to £145,508); the largest contributors to this are costs from social care, short-term health and long-term labour market outcomes. The discounted lifetime cost per death from child maltreatment is estimated at £940,758, comprising health care and lost productivity costs. Our estimates provide the first comprehensive benchmark to quantify the costs of child maltreatment in the UK and the benefits of interventions aimed at reducing or preventing it.

## 1. Introduction

Child maltreatment has increased in developed nations since the 1970s, despite varied policy initiatives aimed at preventing or reducing it (Gilbert et al., 2011). In England and Wales, in the last twenty years there has been a steep rise in the incidence of crimes against children, of child protection registrations, and of children entering care during 2000–16 (Degli Esposti et al., 2019). In particular, over the last decade, in England there has been a substantial increase in the number of children becoming involved in the child protection system, with those subject to child protection investigations rising from 87,700 in 2009/10 to 179,160 in 2018/2019 (Department for Education, 2010 and 2019). More recently, a substantial increase in child maltreatment is feared to be one of the consequences of the ongoing covid-19 pandemic and the associated lockdown policies (Conti, 2020). The total number of serious incident notifications where a child had died or been seriously harmed reported to the Department for Education in England during the first half of 2020-21 increasing by 27% on the same period in 2019-20.^2^

The NSPCC describes child maltreatment as “all forms of physical and/or emotional ill-treatment, sexual abuse, neglect or negligent treatment or commercial or other exploitation, resulting in actual or potential harm to the child’s health, survival, development or dignity in the context of a relationship of responsibility, trust or power”. Child maltreatment has long-lasting effects on mental health, drug and alcohol use, risky sexual behaviour, obesity and crime, with high costs for both the individual and the society (Norman et al., 2012). While there are estimates of the costs of child maltreatment for different countries, such as US (Fang et al., 2012; Peterson et al., 2018) and Australia (McCarthy et al., 2016), scarce is the evidence for the United Kingdom.

The aim of this study is to calculate the first estimates of the lifetime costs per victim of nonfatal and fatal child maltreatment by primary caregivers from a societal perspective in the UK using an incidence-based approach. While previous studies have used only secondary evidence to quantify the costs of child maltreatment (as for example in Fang et al., 2012), we develop a novel approach by combining new estimates from two British cohorts on the medium- and long-term consequences of child maltreatment with secondary evidence from the literature; in this way, our cost estimates are based on a more coherent set of results than those produced in previous studies. More specifically, we assess the costs of child maltreatment in two steps. In a first step, we estimate robust associations of child maltreatment with medium- and long-term outcomes by means of regression analysis, and we address selection issues by controlling for an extensive set of background and socioeconomic conditions; additionally, we perform a formal test for omitted variable bias developed by Oster (2019). For associations with short-term outcomes, we use recent results from the literature, given the lack of primary data. In a second step, we compute the costs over a lifetime horizon, including those related to short- and long-term health, crime, social care, special education, and productivity losses; additionally, we provide extensive sensitivity analyses.

The discounted average lifetime incidence cost of non-fatal child maltreatment by a primary caregiver is estimated at £89,390 (95% uncertainty interval £44,896 to £145,508); the largest contributors to this are costs from social care, short-term health and long-term labour market outcomes. The discounted lifetime cost per death from child maltreatment is estimated at £940,758, comprising health care and lost productivity costs. Our estimates provide the first comprehensive benchmark to quantify the costs of child maltreatment in the UK and the benefits of interventions aimed at reducing or preventing it.

## 2. Literature Review

Our work relates to three different strands of literatures: papers which have studied determinants and consequences of child maltreatment using data from the UK, economic papers on child maltreatment, and papers which have tried to assess its costs. Hence, before conducting our analysis, we performed three literature reviews: the first one of work based on UK data, the second one of papers published in economic journals, and the third one of cost-focused studies. The full list of papers for each of the three reviews performed is presented in Appendix 1.

The first literature review showed that both survey/cohort and administrative data have been used to date for UK studies. The UK survey/cohort data used in the published literature are: the National Child Development Study (NCDS), the Avon Longitudinal Study of Parents and Children (ALSPAC), the Environmental Risk Longitudinal Twin Study (E-Risk), and more recently the UK Biobank study (UKB). For what concerns the use of administrative data: two studies (Woodman et al., 2012, and Chandan et al., 2019) have used primary care records, two studies (González-Izquierdo et al., 2010 and 2014) have used data from the Hospital Episode Statistics, two studies (Mc Grath-Lone et al., 2016 and 2017) have used administrative records on Children Looked After and two very recent studies (Murray et al., 2020a and 2020b) have used data from the Longitudinal Study; additionally, another recent study (Baldwin et al., 2020) has used the Born in Bradford cohort linked with data from the Child Protection System. This review also showed that using directly results from this literature as basis for our cost estimates was problematic, since different definitions of child maltreatment have been adopted (even in studies based on the same data, see Table A1 of the Supplementary Material), and different methodologies and control variables have been used in the various studies. The majority of the studies documented associations between child maltreatment and a variety of outcomes across the lifecycle, ranging from greater prevalence of cardiovascular diseases, multi-morbidities, mental health illness, higher BMI and (premature) mortality, and worse socioeconomic outcomes among children maltreated or placed in out-of-home care (e.g. Chandan et al., 2019; Fahy et al., 2017; Geoffroy et al., 2016; Hanlon et al., 2020; Ho et al., 2020; Soares et al., 2020; Murray et al., 2020, 2021; Power et al., 2015). Other studies examined the determinants of child maltreatment, and noted that individual-, family-, environment-, community-level and also genetic-level factors are all relevant in understanding its aetiology (e.g. Sidebotham et al., 2001, 2002, 2003, 2006; Jaffee et al., 2004; Baldwin et al., 2020) Few other studies (e.g. Collishaw et al., 2007) investigated the intergenerational consequences of childhood abuse, trends in child maltreatment over time (Gonzalez-Izquierdo et al., 2010, 2014), and the patterns of entry and exit from out-of-home care (McGrath-Lone et al., 2015, 2017).

The second literature review showed that few papers have been published in economic journals on the causes and consequences of child maltreatment (see Doyle and Aizer, 2018, for a recent review), with only one paper (Schurer et al., 2019) based on UK data from the 1958 birth cohort. Among the studies with the more robust design, Currie and Tekin (2012) use a siblings and twins fixed effects strategy based on data from the National Longitudinal Study of Adolescent Health (Add Health) to find that maltreatment greatly increases the probability of engaging in crime and that the probability increases with the experience of multiple forms of maltreatment; and Doyle (2007 and 2008) uses the placement tendency of child protection investigators as an instrumental variable to identify causal effects of foster care on long-term outcomes, and finds that children on the margin of placement tend to have better outcomes when remain at home.^3^

The third and last literature review identified 29 cost studies, none of which, however, estimated lifetime costs for UK using an incidence-based approach. The most common objective of the included studies was to estimate the cost of child maltreatment (in general across all types of abuse combined, or for some specific form of abuse); the majority of the studies adopted the health care provider perspective, the hospital perspective or the societal perspective. There are several incidence-based studies from the USA (Fang et al., 2012; Peterson et al., 2018), Europe (Sethi et al., 2013), Germany (Habetha et al., 2012), Australia (McCarthy et al., 2016) and Asia (Fang et al., 2015), but epidemiological data, socioeconomic conditions and costs structure are very different from the UK. Among the few UK studies, one conducted in the UK Royal Liverpool Children’s Hospital (Summers and Molyneaux, 1992) estimates the financial implications of child maltreatment from the hospital perspective; however, the study refers to children hospitalised in 1990, therefore the results are out of date and take a narrow cost perspective. Another UK study has estimated the cost of sexual abuse in the UK using a prevalence approach (Saied-Tessier, 2014). Hence, to date, no comprehensive cost analysis of child maltreatment for UK exists, despite the high and increasing prevalence of this phenomenon. Our study fills this significant gap in the literature.

## 3. Data and Methods

As mentioned in the introduction, we conduct our analysis in two steps: in a first step, we obtain the association between child maltreatment and a variety of short-, medium- and long-term outcomes; in a second step, we compute the costs. In the following we describe the data and the methods used in each of these steps in turn.

### 3.1 Data and methods for the regression analysis

We use two datasets to estimate the medium- and long-term effects of child maltreatment:^4^ the National Child Development Study (NCDS), i.e. the 1958 British birth cohort, and the English Longitudinal Study of Ageing (ELSA). The NCDS (Power et al., 2005) follows the lives of over 17,000 people born in England, Scotland and Wales in a single week of 1958; it is the dataset which has been mostly used in the UK literature to examine the effects of child maltreatment. The ELSA (Steptoe et al., 2012) is a longitudinal survey of ageing and quality of life among people aged 50 and above, which began in 2002.

In the NCDS, both prospective and retrospective measures of child maltreatment are available, and both have been used in the past literature. Few papers (see for example Newbury et al., 2018 among the more recent) compare prospective and retrospective measures: in general, both have shortcomings and both should be used, since they are likely to capture non-overlapping groups of maltreated children. However, the prospective measures in the NCDS are only related to neglect, and consist in teachers assessments reported at three points in time (ages 7, 11 and 16 of the cohort member) on particular aspects of the child (‘looks undernourished, scruffy or dirty’), on the parental level of interest and aspiration for the child’s education, and on the amount of time the parents spends with the child; no prospective measures are collected in the NCDS on any form of abuse. The retrospective measures, instead, have been asked in the biomedical sweep to approximately 9,400 cohort members when they were 44/45 years of age, and they directly refer to both neglect and physical, emotional, and sexual abuse; the precise wording used is detailed in Appendix 2 of the Supplementary Material. After consultation with experts in the field, we opted for the retrospective measures because they were directly related to different types of child maltreatment, while the prospective ones were only indirectly related to neglect (as such also potentially subject to interpretational biases); and we constructed a ‘global’ measure, which includes any type of maltreatment by a primary caregiver by age 16 (reported retrospectively at age 44/45), to both overcome the lack of costing data disaggregated by the type of child maltreatment, and also to avoid potential double-counting. In the ELSA, only retrospective measures of neglect and physical abuse are available, which were asked in the life history module in the third wave (when the subjects were age 53 or above); to the best of our knowledge, they have not been used in any other published paper. Like for the NCDS, we combined them into a ‘global’ measure, indicating whether the individual reports to have been neglected or physically abused before age 16. The summary statistics (Appendix 2) show that 12.4% of the NCDS respondents report to have been neglected in childhood (the corresponding figure for ELSA is 3.6%, see Tables 2L and 2M in Appendix 2), another 12.4% report to have received a form of emotional abuse, 6% report to have experienced physical abuse (the corresponding figure for ELSA is 3.5%, see Table 2N), and 1.5% report to have experienced sexual abuse. Combining them all together into our ‘global’ measure, 20.6% of the NCDS respondents has been victim of some form of child maltreatment: this is very similar to the figure in Radford et al. (2013) for UK, who report that 24.5% of young adults have experienced a form of child maltreatment at least once during childhood, in a random representative sample interviewed in 2009.

To study the consequences of child maltreatment, we then selected the following medium- and long-term outcomes to be used in the regression analysis, on the basis of the literature review: obesity, hypertension, diabetes, cancer, any diagnosed mental health problem, anxiety, depression, heavy drinking (consuming two or more alcoholic drinks/day), smoking, heavy smoking (25 cigarettes or more/day), employment, net and gross earnings and disability benefits. The vast majority of the outcomes was surveyed in both the NCDS and in the ELSA, and we put great care in making them comparable to the extent possible.

Clearly, the key challenge in the empirical analysis arises from the fact that parents maltreating their children are not a random sample of the population, hence the effect of maltreatment needs to be disentangled from that of other unobserved characteristics which are also correlated with the outcomes of interest. Given the unavailability of information on siblings or twins in our data, or of any credible instrument, our approach consists in estimating different specifications including incrementally more controls for background and socioeconomic conditions, pregnancy and birth circumstances, and other adverse early experiences, and to check the robustness of our results to the inclusion of such controls (a similar methodology has been used in Schurer et al., 2019); we also formally implement a test for omitted variable bias proposed by Oster (2019) which corroborates the robustness of our results. For the cost analysis, we use the estimates from the most conservative specifications.

We use linear probability models throughout for computational convenience; logistic regression models (available upon request) yielded qualitatively similar results. We estimate four different specifications, incrementally adding different sets of controls. The first specification only includes a basic set of demographic controls (binary indicators for gender, ethnicity and interview date); the second specification adds as controls an extended set of demographics recorded at birth (the social class of the husband and of the father of the mother, parity, birthweight, whether the mother stayed in school beyond the minimum school leaving age, marital status, smoking in pregnancy, mother’s and father’s age at the child’s birth, working during pregnancy, weight before pregnancy, mother’s height, whether the mother had any antenatal visit in the first trimester of pregnancy, and the number of people per room); the third specification controls for other early adverse childhood experiences, collected in the age 6 sweep (whether the child lived with both natural parents, whether there was any family contacts with the probation officer, whether the parents divorced, whether a parent was alcoholic, whether the death of the father or of the mother occurred, whether there was any domestic tension, whether there were financial or housing difficulties, and the number of times the family moved since the birth of the child); the fourth specification further controls for an extended set of early adverse childhood experiences, reported retrospectively in the biomedical sweep (whether the mother suffered from nervous or emotional trouble, whether the mother had trouble drinking, whether there was some or a lot of conflict and tension in the house, whether the child grew up in poverty or in financial hardship).

### 3.2 Data and methods for the cost analysis

In the second step of our approach, we use the results from the regression analysis carried out in the first step, supplemented with the best published evidence, to develop our cost estimates for the medium- and long-term outcomes: anxiety, depression, smoking, alcohol abuse, productivity losses, special educational and social care needs, and police, court and penal services. We prefer the NCDS results over the ELSA results (apart from heavy drinking), since the former incorporates more types of child maltreatment in the global measure; as mentioned, we use the estimates from the specification with the extended set of controls (including indicators for several other early adversities, such as parental separation or death), so that our results can be considered conservative. Then, we combine the medium- and long-term costs based on our novel NCDS and ELSA results with published UK figures for short-term costs caused by unplanned hospital admissions, maltreatment or violence-related injuries and emergency treatment of hyperkinetic, conduct or emotional disorders; we could not directly estimate from the data the association of child maltreatment with short-term outcomes because we did not have details on the exact age at which child maltreatment occurred (only that it occurred before age 16). The list of all the outcomes included in the computation of the costs is in Table 1. The analysis excludes costs associated with obesity, hypertension, type 2 diabetes, and cancer, as these were not significantly related to the global measure of child maltreatment in our regression results (see Table 2). We did, however, include costs of other conditions that are related to depression, anxiety, smoking and alcohol abuse where these are included in published cost estimates (e.g., the cost of developing lung cancer among smokers is included in the smoking-related costs). The analysis also excludes costs of days off work among those who are employed as direct consequence of child maltreatment, as this variable was not available in the datasets we used; however, these costs were included in the calculations of long-term health-related costs where they were included in published cost estimates (e.g., absenteeism due to depression). To avoid double counting, when productivity losses from long-term health-related problems were included, the productivity losses due to reduced employment associated with child maltreatment were removed. We also found that child maltreatment was not associated with lower wages among those who were employed, so we did not include this cost. Also, we did not evaluate the association of child maltreatment with education attainment directly, but we included special education costs associated with child maltreatment and accounted for education effects indirectly in the calculation of productivity losses.

**Table 1:** Outcomes included in the calculation of the costs of child maltreatment.

|  |
| --- |
| **Health** |
| **Short-term** |
| Unplanned hospital admissions for injuries: MVR rate for different year groups and admission expenses |
| Stress-induced disorders in children: Hyperkinetic, Conduct and Emotional |
| **Long-term** |
| Depression* |
| Anxiety* |
| Smoking* |
| Alcohol (heavy drinking at age 50 or more)* |
| **Criminal justice** |
| Court proceeding and convictions expenses |
| Sex offenders treatment programmes |
| Proportion of cases ending in court |
| Proportion of cases ending in convictions |
| Proportion of cases of sexual abuse ending in sex offenders treatment programmes |
| **Social care** |
| Foster care expenses |
| Local Authority care expenses |
| Child Protection Plan involvement |
| Monthly childcare support |
| Proportion newly looked after children receiving intervention in foster care |
| **Productivity** |
| Special education |
| Employment at different ages* |
| Receipt of disability benefits* |

**Note:** \*The outcomes depression, anxiety, smoking, alcohol, employment and receipt of disability benefits are based on the regression analysis of NCDS and ELSA; we combine this regression analysis with costs taken from published source. For the remaining outcomes, the computation of the costs is entirely based on secondary evidence from the literature.

**Table 2:**
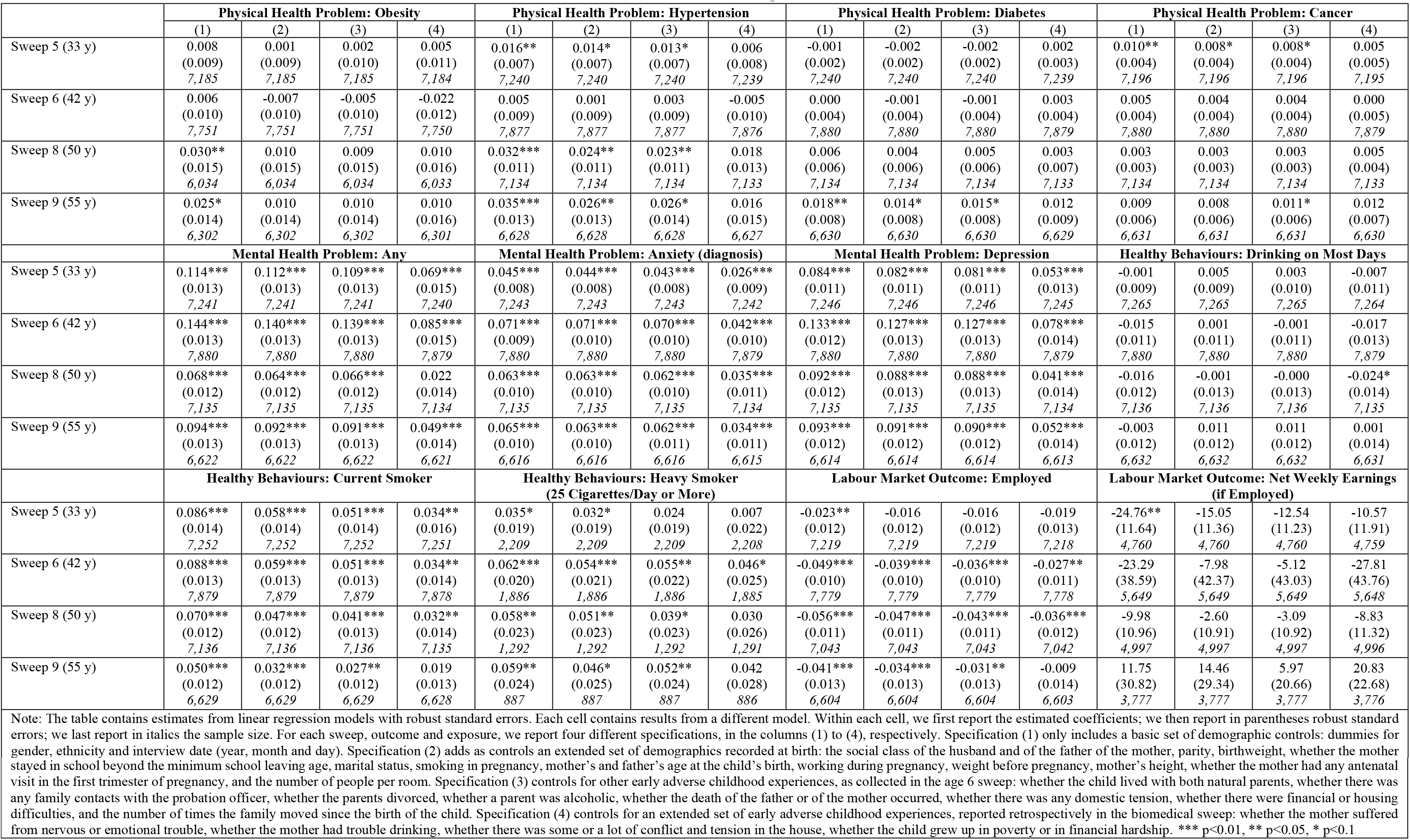
NCDS Results of the Effects of the global measure of CM.

All costs were measured in GBP and adjusted to 2015 prices. We assumed that the average age at which maltreatment starts is 6 years,^5^ and that incidents are assumed to occur up to age 18 (DfE, 2016). Costs are presented in present value terms: future costs for the ages 6-30 years, 31-74 years and over 75 years are discounted using annual rates of 3.5%, 3% and 2.5%, respectively. Costs related to productivity losses are assumed to end at 67 years.

As explained above, given that in both the NCDS and ELSA the exact age of maltreatment was not specified (only the fact that it happened before age 16), we based our calculations of short-term costs on published figures. Secondary evidence was identified via searches of several databases including PubMed, the NHS Economic Evaluations Database, EconLit, Google Scholar and Google. For each cost category, we used published estimates from previous cost of illness studies where possible. We used data that were specific to the UK as opposed to countries outside of the UK because health care, social care, education and criminal justice systems vary between countries and costs in other countries are unlikely to apply to the UK. The only exception to this was in sensitivity analyses, where we applied costs of child maltreatment calculated for the USA to the UK to compare differences between the two studies. Costs vary over time and we used recent data whenever possible, though as noted below several of the data sources are dated.

We did not evaluate costs by type and severity of child maltreatment and instead opted for an overall maltreatment estimate. One reason is that many of the studies used to provide inputs into the calculations did not distinguish by type of maltreatment, so disaggregating costs would be difficult. In the econometric analysis, it was not possible to analyse the impact of different types of maltreatment on long-term health outcomes and labour market outcomes, since small numbers of cases for some types of maltreatment meant the analysis was underpowered. Another issue is that there may be overlap between different types of maltreatment, making it difficult to attribute costs to individual types.

Although our aim was to estimate the lifetime cost per victim of child maltreatment, for several of the cost components data were only available at the aggregate level across all victims. Where costs were available at the aggregate level across all victims only, they were divided by an estimate of the number of new cases of maltreatment per year. Several of the cost components required longitudinal data on the number of events for new cases of child maltreatment in a single year in order to accurately calculate incidence-based costs. Where data was insufficient to calculate accurate incidence-based costs, proxies for event frequency were created under steady-state assumptions. Using this approach, the number of events in a single year across all victims of child maltreatment is a proxy for all events over time among new victims in that year. This assumption requires that the number of events remains fairly constant over time, which may be problematic for some of the cost components considered.

A summary of the cost components included in the analysis is presented in list form in Table 4. We first estimated short-term health-related costs in the form of unplanned hospital admissions for maltreatment or violence-related injuries by using incidence rates as a proxy for total admissions (ordinary and day case) for new victims (González-Izquierdo et al., 2014): we applied the unplanned admission rate for (diagnosed) maltreatment or violence related injuries in England in 2011 by age group to the mid-year UK population estimates to assess the number of unplanned admissions in 2015 in UK. Combined with the national average unit cost of non-elective admissions for paediatric injuries (DoH, 2015) and divided by an estimate of maltreatment occurrence, those figures yielded average costs of £120 per victim. Additionally, we calculated the prevalence of mental health problems for children who were maltreated (Table A2, based on Meltzer et al., 2003). Combined with costs reported by Snell et al. (2013), multiplied by the probability of each disorder and summed across different disorder types, these figures yielded a discounted cost of £11,453 per victim. Supplemented by evidence on increased criminal justice costs associated with conduct disorders (Scott et al., 2001), those generated a total of £18,553 in short-term mental health costs per victim.

For long-term health-related costs, we firstly estimated the mean incremental lifetime costs of anxiety and depression as the product of their average costs per year as indicated by McCrone et al. (2008) and Fineberg et al. (2010) (Table 5) and the marginal effect of child maltreatment on the probability of the respective outcome at each year of age (linearly extrapolated where missing). Costs were estimated to be £954 per year per victim for anxiety and £5,145 for depression. Similarly, combining the NCDS results with estimated social annual costs and smoking population data by Action on Smoking and Health (ASH 2016), the discounted mean incremental lifetime cost of smoking was estimated at £528 per victim. Equally, the discounted lifetime cost of alcohol misuse associated with maltreatment (assumed to begin at age 50 and remain constant), calculated with cost figures by the Cabinet Office (2003) and ELSA estimates, was £537.

For criminal justice system costs, the proportion of sexual offence cases resulting in court proceedings and convictions — derived from police records — was applied to the total number of relevant criminal cases in 2015 and used as a proxy for the lifetime number of offences from new maltreatment cases. Multiplying resulting figures by the unit cost of court proceedings, convictions, and sex offender treatment programmes yielded an average cost of £4,316 per victim. Next, we estimated social care costs, multiplying the number of children entering child protection plans or registers in 2015 (minus the number of re-registrations, DfE 2015) by the fixed and ongoing costs of protection initiatives. We further estimated the cost of children in foster care or local authority homes using the English data for all newly looked after children (DfE, 2015). This was based on the proportion of looked after children due to maltreatment in England and Wales (61% and 66% respectively) in 2016 (NSPCC, 2016) and on the incidence of child maltreatment in 2015 (DfE, 2016). In absence of data, costs of being placed for adoption, parents or community placement, were not considered. The actual costs for child social care thus likely exceed the estimated total of £2,360,129,680 or £38,132 average cost per victim (Tables 4 and A3). We excluded costs from drug use, divorce and disability for lack of data, as well as intangible costs, such as emotional suffering, which are difficult to quantify.

For productivity costs due to educational losses, we used the incremental effect of child maltreatment on special education:^6^ the undiscounted cost per maltreated child in receipt of educational support from age 6 to 16 was calculated as £7,068 per victim, using the DCSF (2009) Schools Census. Lastly, productivity losses due to reduced employment were calculated as the product of earnings and the marginal effect of maltreatment on employment, assumed to be zero before 33 years and after 55 years, and linearly interpolated in between. Losses were valued by age-specific earnings before deduction of taxes in 2013-14 inflated to 2015 prices (ONS, 2014), yielding an average discounted lifetime productivity loss of £14,037. The central estimate of the mean total lifetime costs of non-fatal child maltreatment per victim was calculated by summing per-victim costs across all components.

We performed extensive sensitivity analyses to deal with the uncertainty in our estimates. Probabilistic sensitivity analysis (PSA) was used to calculate 95% uncertainty intervals at the 2.5th and 97.5th percentiles (Table A8). For each of the 1000 simulations, all parameters with assigned probability distributions were selected at random from beta distributions for uncertainty in the probabilities, and gamma or triangular distributions for uncertainty in costs (Briggs et al., 2006). Univariate deterministic sensitivity analysis was used to explore the sensitivity of the central estimates to individual parameter values related to: the discount rate, unplanned injury-related admissions, health care and criminal justice system costs, the impacts of child maltreatment, the number of cases ending in court proceedings, the unit costs of child protection plans, monthly ongoing support, special education, and the wages. In Table 5 we summarise the assumptions of the univariate sensitivity analysis and we report the central values and the changed values for each parameter; for example we assumed a base case discount rate of 3.5% up to 30 years and varied it between 0 and 5%. For all outcomes, we further explored the impact of using US estimates (Fang et al., 2012). Changes producing total lifetime costs £10,000 higher or lower than the central estimate were judged substantive.

Lastly, the lifetime cost per victim of fatal child maltreatment was estimated in two components, following Fang et al. (2012). Firstly, health care costs from fatal injuries were calculated based on the mean cost of £13,863 per fatal blunt trauma and £5,408 per penetrating trauma injuries in the UK, including the costs of hospital transport, stays, and procedures (Christensen et al., 2008a and 2008b). The lower value was used for the central estimate, the higher for sensitivity analysis. Second, lifetime costs of lost productivity were calculated using the human capital approach combining mean annual earnings — discounted to present value terms and inflated assuming a 2% constant annual increase in earnings — and employment rates by age (ONS, 2016). Those were summed across the lifetime and assumed to represent mean lifetime productivity — lost with an early fatality. For these calculations, probabilistic sensitivity analysis was not possible because probability distributions could not be assigned to key parameters, so univariate deterministic sensitivity analysis was undertaken, varying the discount rate, health care costs, assumptions concerning the lost productivity and criminal justice costs.

## 4. Results

The main NCDS results on the associations between child maltreatment (our ‘global’ measure) and medium- and long-term outcomes are reported in Table 2;^7^ secondary results from the age 23 sweep of the NCDS and from the ELSA are reported in Tables A5, A6, A7 in the Supplementary Material. Our preferred results are those for the most controlled specification (column 4 in Table 2, and column 2 in Tables A6 and A7). They show that having experienced maltreatment in childhood is associated with worse mental health outcomes (in particular anxiety and depression, with increases ranging from 2.6 to 7.8 percentage points in the NCDS and from 1.0 to 6.9 p.p. in the ELSA), and with a higher probability of being a current smoker (3.2-3.4 p.p. increase in the NCDS) and a heavy drinker (2.9 p.p. increase in the ELSA) in adulthood. These results are present since early (30s) until late adulthood (mid 50s or 70s, depending on the data), are remarkably similar in the two cohorts, and are robust to the inclusion of an extended set of controls, including an extensive set of adverse early childhood experiences (to try and isolate the effect of being maltreated versus other early adversities). The strong results on mental health are in line with other studies, also based on administrative data: for example, Chandan et al. (2019) find that having been maltreated doubles the risk of developing mental ill health, using primary care electronic health records from the UK.

The NCDS results also show a 2.7 and 3.6 percentage point reduction in the probability of being in paid employment at ages 42 and 50, respectively, for victims of child maltreatment. The ELSA results further show an increase in weekly disability-related benefits receipt by £36 for the victims of abuse (Table A7); however, these were not included in the cost analysis, since they are transfer payments (Luce et al., 1996). On the other hand, we are unable to detect any robust associations with several physical health measures, such as obesity, hypertension, diabetes and cancer, and with earnings (conditional on employment). These results are not entirely consistent with some other studies based on UK data (even based on the same NCDS data we use), which instead detected significant associations with these outcomes, although comparisons are significantly hindered by differences in samples, methods and choice of control variables; for example, none of the previous studies controls for other forms of child adversity (like we do in our more controlled specification), which are likely to co-occur with child maltreatment and so potentially bias estimates of its impacts.

Despite controlling for an extensive set of covariates, there is still the possibility that our estimates might be biased by omitted variables. Two facts however reassure us against that. First, previous results on child maltreatment from studies using robust designs: Currie and Tekin (2012) notice (p.528) that the estimated effects from siblings fixed effects models are remarkably similar to the OLS results; in Doyle (2008) the 2SLS coefficients are much bigger than the OLS coefficients, which would suggest that – if anything - our estimates are very conservative and we are estimating a lower bound. Second, to formally assess the robustness of our results, we have performed the test developed by Oster (2019), to examine the extent to which omitted variables could bias the relationship between child maltreatment measures and our set of outcomes. This test uses movements in the coefficient of interest and in the R2 after adding observable controls to learn about the likely impact of the unobservables. The results are shown in Table 3 for the NCDS results: the estimates of the coefficients of proportionality (Delta) in general are above one, suggesting that unobservables would have to be more important than observables for the coefficients to be zero – an unlikely occurrence, given the extensive set of results we include in our most controlled specification.

**Table 3:**
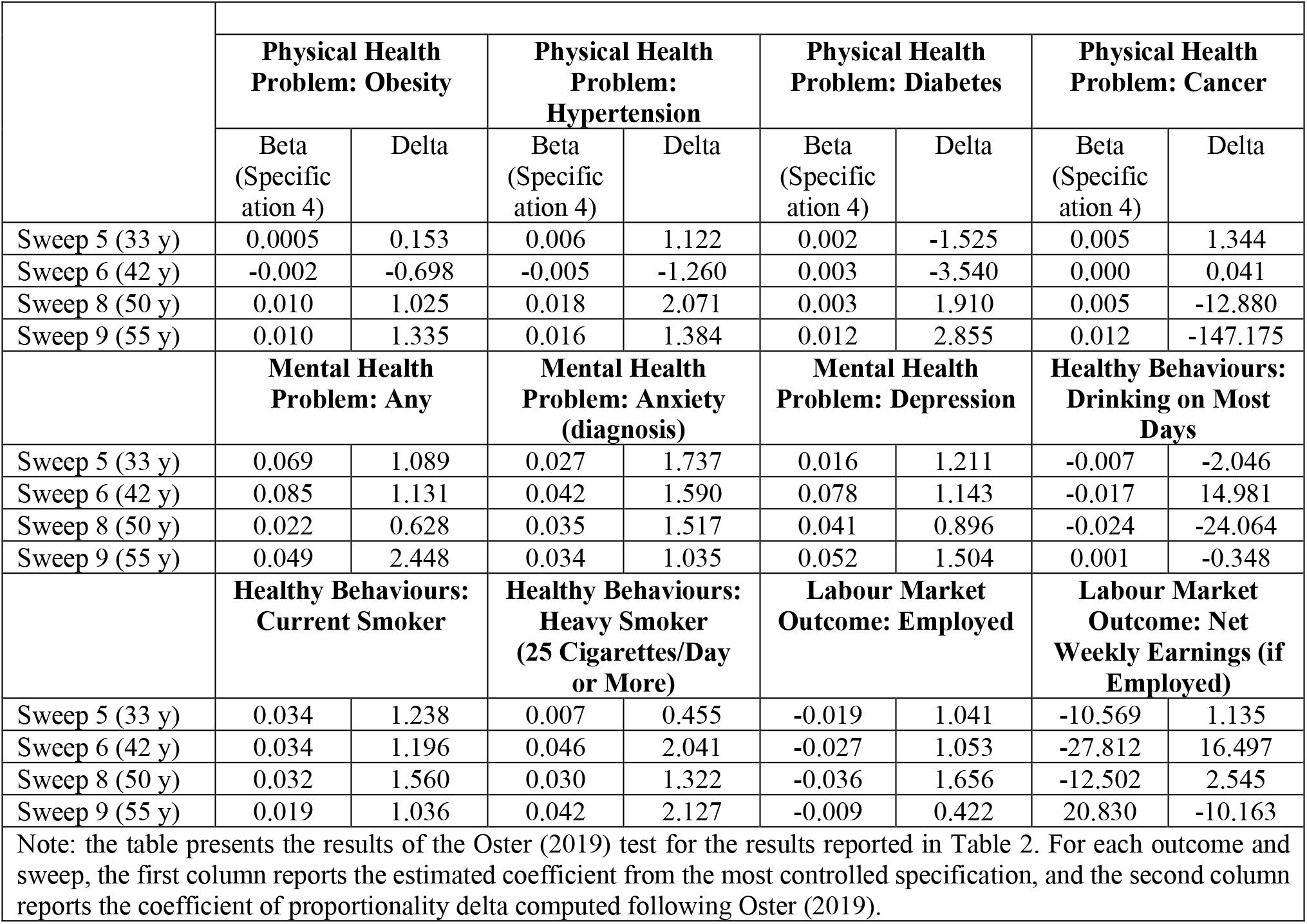
Results of the Oster (2019) Test.

Using the results of the regression analysis, the marginal effect of child maltreatment on the probability of experiencing an outcome (e.g. anxiety, depression, smoking, drinking alcohol etc) at each year of age are applied to the unit average cost associated to each outcome. For example, if child maltreatment increases by 5.3% the probability of experiencing depression at age 33 and 7.8% at age 42, assuming a linear relation between the marginal effects at different ages, we apply the unit cost per year to the increased probability to estimate the incremental cost to treat depression that is associated to child maltreatment. The same is done for all the statistically significant results.

Combining the regression results with secondary cost data, we calculate a discounted average lifetime cost of non-fatal child maltreatment per victim of £89,390 (95% uncertainty interval £44,896 to £145,508). The main contributors are social care, short-term health-related, and reduced employment costs (Table 4). In Figure 1a we summarise graphically the distribution of values and we calculate the probability that the total cost was greater than pre-specified values as the proportion of the simulations greater than values £0 to £200,000 (Figure 1a). The intervals are wide, reflecting uncertainty in the parameters: there is a 96% probability that the total cost is greater than £50,000, a 34% probability that is greater than £100,000, and a 3% probability that is greater than £150,000 (Figures 1a, 1b).

**Table 4:** Discounted lifetime costs per victim of non-fatal child maltreatment.

|  | **Value (£)** | **95% Uncertainty Interval** | **% Total** |
| --- | --- | --- | --- |
| Unplanned hospital admissions for injuries | 120 | (83, 141) | <1 |
| Short-term mental health problems | 18,553 | (9,758, 29,833) | 21 |
| Short-term health-related costs | 18,673 | (9,841, 29,974) | 21 |
| Anxiety | 954 | (311, 2,094) | 1 |
| Depression | 5,145 | (1,782, 10,740) | 6 |
| Smoking | 528 | (100, 1,461) | 1 |
| Alcohol abuse | 537 | (148, 1,262) | 1 |
| Long-term health-related costs | 7,164 | (2,341, 15,558) | 8 |
| Criminal justice system costs | 4,316 | (2,509, 6,165) | 5 |
| Social care costs | 38,132 | (22,679, 53,346) | 43 |
| Special education costs | 7,068 | (2,162, 14,455) | 8 |
| Reduced employment | 14,037 | (5,364, 26,010) | 16 |
| Total | 89,390 | (44,896, 145,508) | 100 |

**Note:** All costs are discounted and in 2015 UK£. The outcomes anxiety, depression, smoking, alcohol abuse, and employment, are based on the regression analysis of NCDS and ELSA.

**Figure 1.**
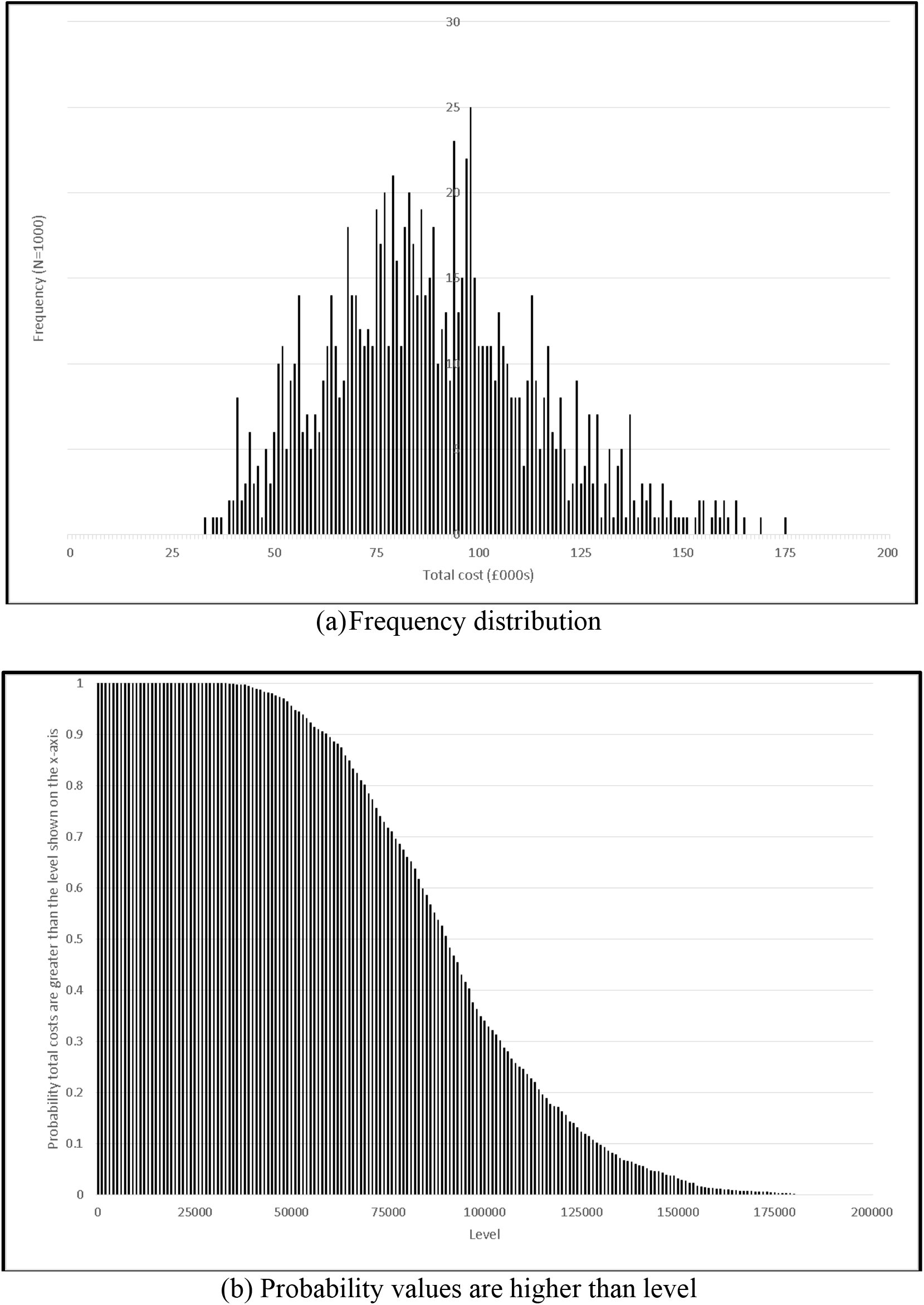
Discounted lifetime costs per victim of non-fatal child maltreatment: distribution of values from probabilistic sensitivity analysis.

Deterministic sensitivity analysis (Table 5) suggests that the central estimate is sensitive to: the discount rate (values recommended by HM Treasury were used for final results), the marginal effects of child maltreatment on mental health problems, and to changes in assumptions on social care and employment costs (Table 5); the central estimate remains stable in the other cases. The discounted lifetime cost of fatal child maltreatment is calculated as £940,758, comprising £5,408 health care costs and £935,350 lost productivity costs (Table 6). As for the cost of non-fatal child maltreatment, these results are sensitive to the discount rate and assumptions concerning productivity losses (Table 7). While there is no comparable study using UK data, direct comparisons with the Fang et al. (2012) approach based on US data are complicated by systemic cross-country differences, as well as differences in methodology. Estimated lifetime costs per victim of non-fatal child maltreatment are thus noticeably different (£89,930 versus US$210,012); estimated costs per death from child maltreatment however are broadly comparable (£940,758, versus US$1,272,900). Differences are even more marked with Peterson et al. (2018), who estimate lifetime cost per victim of non-fatal maltreatment at $830,928 (2015 USD) and per-victim cost of fatal child maltreatment at $16.6 million. However, their calculations also incorporate victim and intangible community costs, suggesting that the UK costs we have calculated are likely a lower bound of the full costs of child maltreatment.

**Table 5:** Discounted lifetime costs per victim of non-fatal child maltreatment: univariate deterministic sensitivity analysis.

|  |  | **Value (£)** |
| --- | --- | --- |
| **Central estimate** |  | 89,390 |
| **Central estimate assumption** | **Changed to (all else equal)** |  |
| Discount rate |  |  |
| Years 0 to 30 = 3.5% per annum; years 31 to 75 = 3% per annum; years 76 to 125 = 2.5% per annum | All years = 0% per annum | 148,747 |
| Years 0 to 30 = 3.5% per annum; years 31 to 75 = 3% per annum; years 76 to 125 = 2.5% per annum | All years = 5% per annum | 75,884 |
| Unplanned hospital admissions for injuries |  |  |
| 8,685 unplanned admissions across all victims | Each victim has one unplanned admission; 31.2% have a second admission | 90,389 |
| 8,685 unplanned admissions across all victims | Each victim has one unplanned admission; 31.2% have a second admission; each admission is associated with 11.5 outpatient visits and 11.5 GP visits | 92,758 |
| Short-term mental health problems |  |  |
| Annual cost per victim with mental health disorder = £1,351 to £3605 depending on disorder | Annual cost per victim with mental health disorder doubles | 100,843 |
| Marginal effect of child maltreatment on probability of mental health problems = +0.061 to +0.345 depending on age and type of disorder | Marginal effect of child maltreatment on probability of mental health problems = +0.05 | 75,657 |
| Marginal effect of child maltreatment on probability of mental health problems = +0.061 to +0.345 depending on age and type of disorder | Marginal effect of child maltreatment on probability of mental health problems = +0.5 | 119,035 |
| Annual criminal justice costs for children with conduct disorder = £3,436 | Annual criminal justice costs for children with conduct disorder doubles | 96,490 |
| Short-term health-related costs |  |  |
| Total short-term costs as in base case | Total short-term health care costs as in Fang et al., updated to 2014/15 UK£ (=£24,167) | 94,884 |
| Long-term health-related costs |  |  |
| Marginal effect of child maltreatment on all types of long-term health care costs = +0.029 to +0.077 in effective age ranges depending on age and condition | Marginal effect of child maltreatment on all conditions and ages (maintaining original effective age ranges) = +0.1 | 97,571 |
| Marginal effect of child maltreatment on all types of long-term health care costs = +0.029 to +0.077 in effective age ranges depending on age and condition | Marginal effect of child maltreatment on all conditions and ages (maintaining original effective age ranges) = +0.2 | 112,916 |
| Marginal effect of child maltreatment on all types of long-term health care costs = +0.029 to +0.077 in effective age ranges depending on age and condition | Marginal effects of child maltreatment at youngest effective age extrapolated to 18, and at oldest effective age extrapolated to 100 | 93,469 |
| Total long-term costs as in base case | Total long-term health care costs as in Fang et al., updated to 2014/15 UK£ (=£7,795) | 90,021 |
| Criminal justice system costs incurred by perpetrators |  |  |
| 17.6% cases end in court proceedings and 10.7% end in convictions | All cases end in court proceedings and convictions | 115,045 |
| Criminal justice costs as in base case | Criminal justice cost as in Fang et al., updated to 2014/15 UK£ (=£3,533) | 88,607 |
| Social care costs |  |  |
| Unit cost of child protection plan = £5,321; monthly child care support cost = £328 | Unit cost of child protection plans and monthly child care support costs double | 96,900 |
| Child social care costs as in base case | Child social care costs as in Fang et al., updated to 2014/15 UK£ (= £5,721) | 56,979 |
| Special education costs |  |  |
| Unit cost of special educational support per victim = £3,740 per annum | Unit cost of special educational needs provision doubles | 96,459 |
| Marginal effect of child maltreatment on receipt of special education support = 0.22 | Marginal effect of child maltreatment on receipt of special education support = 0.11 | 85,863 |
| Marginal effect of child maltreatment on receipt of special education support = 0.22 | Marginal effect of child maltreatment on receipt of special education support = 0.33 | 92,946 |
| Reduced employment |  |  |
| Earnings if employed = national average earnings by age group | Earnings if employed = 75% of the national average earnings by age group | 85,881 |
| Earnings if employed = national average earnings by age group | Earnings if employed = 50% of the national average earnings by age group | 82,372 |
| Annual increase in wages 2% | Annual increase in wages 1% | 84,876 |
| Annual increase in wages 2% | Annual increase in wages 3% | 96,008 |
| Marginal effect of child maltreatment on not being employed = 0 at age 33, -0.027 at age 42, -0.035 at age 50 and 0 at age 55, with linear interpolation | Marginal effect of child maltreatment on not being employed at ages 42 and 50 = -0.05, all else equal | 98,349 |
| Marginal effect of child maltreatment on not being employed = 0 at age 33, -0.027 at age 42, -0.035 at age 50 and 0 at age 55, with linear interpolation | Marginal effect of child maltreatment on not being employed at ages 42 and 50 = -0.1, all else equal | 121,344 |
| Marginal effect of child maltreatment on not being employed = 0 at age 33, -0.027 at age 42, -0.035 at age 50 and 0 at age 55, with linear interpolation | Marginal effect of child maltreatment on not being employed at ages 42 and 50 = -0.2, all else equal | 167,334 |
| Marginal effect of child maltreatment on not being employed = 0 at age 33, -0.027 at age 42, -0.035 at age 50 and 0 at age 55, with linear interpolation | Marginal effects of child maltreatment at age 42 (= -0.027) extended to age 18, and at age 50 (= -0.035) extended to age 67 | 113,515 |
| Cost of reduced employment as in base case | Cost of reduced employment as in Fang et al., updated to 2014/15 UK£ (=£4,360 per annum from ages 18-64 inclusive, £0 otherwise) | 150,001 |

**Note:** All costs are in 2015 UK£ and discounted unless indicated otherwise. GP = general practitioner.

**Table 6:** Discounted lifetime costs per death from child maltreatment.

|  | Value (£) | % Total |
| --- | --- | --- |
| Health care costs | 5,408 | 1 |
| Lost productivity | 935,350 | 99 |
| Total | 940,758 | 100 |

**Note:** All costs are discounted and in 2015 UK£.

**Table 7:** Discounted lifetime costs per death from child maltreatment: univariate deterministic sensitivity analysis.

|  |  | Value (£) |
| --- | --- | --- |
| **Central estimate** |  | 940,758 |
| **Central estimate assumption** | **Changed to (all else equal)** |  |
| **Discount rate** |  |  |
| Years 0 to 30 = 3.5% per annum; years 31 to 75 = 3% per annum; years 76 to 125 = 2.5% per annum | All years = 0% per annum | 2,994,842 |
| Years 0 to 30 = 3.5% per annum; years 31 to 75 = 3% per annum; years 76 to 125 = 2.5% per annum | All years = 5% per annum | 504,483 |
| Health care costs |  |  |
| Health care costs associated with fatal injury = £5,408 | Health care costs associated with fatal injury = £13,863 | 949,213 |
| Lost productivity |  |  |
| Earnings if employed = national average earnings by age group | Earnings if employed = 75% of the national average earnings by age group | 706,921 |
| Earnings if employed = national average earnings by age group | Earnings if employed = 50% of the national average earnings by age group | 473,083 |
| Employment rate = national average rates by age group | Employment rate = 75% of the national average rates by age group | 706,921 |
| Employment rate = national average rates by age group | Employment rate = 50% of the national average rates by age group | 473,083 |
| Annual increase in wages = 2% | Annual increase in wages = 1% | 663,419 |
| Annual increase in wages = 2% | Annual increase in wages = 3% | 1,348,863 |
| Criminal justice system costs |  |  |
| Not included | Assume all cases result in a court proceedings and a conviction | 972,867 |

**Note:** All costs are discounted and in 2015 UK£.

## 5. Conclusions

This paper has provided the first estimate of the lifetime costs of child maltreatment in UK. We have combined novel regression analysis to estimate associations between child maltreatment and several medium- and long-term health and socioeconomic outcomes, using rich data from the National Child Development Study (the 1958 British birth cohort) and the English Longitudinal Study of Ageing, with published results on short-term outcomes and secondary sources for the costs. We have computed the discounted average lifetime incidence cost of non-fatal child maltreatment by a primary caregiver at £89,390 (95% uncertainty interval £44,896 to £145,508); the largest contributors to this are costs from social care, short-term health and long-term labour market outcomes. The discounted lifetime cost per death from child maltreatment is estimated at £940,758, comprising health care and lost productivity costs.

Our paper has some noticeable key strengths, such as the inclusion of a wide range of cost components (several based on novel regression analysis), and the extensive sensitivity analysis undertaken to evaluate the impact of uncertainty. However, while our estimates are conservative and based on the best possible evidence, they also have limitations. First, some outcomes found relevant in previous studies (such as drug use) were not available in our data; second, the retrospective measures of child maltreatment we used relied on respondent recall; third, our data did not allow us to use some designs such as sibling fixed effects or instrumental variables; fourth, data shortcomings caused uncertainty regarding specific costs and new cases of child maltreatment per year. The sensitivity analyses showed that the estimates for non-fatal child maltreatment are especially sensitive to its impacts on short-term health-related costs, for which there was substantial uncertainty. For all other outcomes, any parameter uncertainty did not affect lifetime costs or the central estimate appreciably.

Several extensions of this work are possible for future research: to include child maltreatment by non-primary caregivers, to calculate the costs of different types of child maltreatment and to use richer data and improved methodological approaches. For the time being, our figures provide the first important indication of how much money the British society is spending on any case of child maltreatment, and consequently, the amount saved if it were prevented. These figures can inform economic evaluations of child maltreatment intervention or prevention initiatives, as well as retrospective analysis of their cost-effectiveness; they can be included in estimates of the costs of the increase in cases of child maltreatment as result of the coronavirus pandemic and associated lockdown restrictions;^8^ and in general they can shape a wider narrative on the significant burden child maltreatment poses on society.

## Data Availability

The data used in the paper are available to interested researchers at the UK Data Archive.

## Acknowledgements

This work was supported by the National Society for the Prevention of Cruelty to Children (NSPCC). EP is supported by the NIHR Collaboration for Leadership in Applied Health Research at Barts Health NHS Foundation Trust (NIHR ARC North Thames). The views expressed are those of the authors and not necessarily those of the NHS, the NIHR or the Department of Health and Social Care. GC was partly supported by the European Union’s Horizon 2020 research and innovation programme (grant agreement No. 633595 DynaHEALTH) and by the European Research Council under the European Union’s Horizon 2020 research and innovation programme (grant agreement No. 819752 DEVORHBIOSHIP – ERC-2018-COG). This report would not have been possible without the input of the advisory group: a huge thank you to Helen Fisher, John Devaney, Chris Cuthbert, Haroon Chowdry, Andrew James, Jon Brown, Alan Wardle, Pam Miller, and Sonja Jütte for their time and input.

## Supplementary Material for “The Economic Costs of Child Maltreatment in UK”

### Appendix 1: Literature review information

The following papers have been reviewed for the respective categories listed below.

#### Studies of Child Maltreatment based on UK data

- Arseneault L, Cannon M, Fisher HL, Polanczyk G, Moffitt TE, Caspi A (2011) Childhood trauma and children’s emerging psychotic symptoms: a genetically sensitive longitudinal cohort study. *American Journal of Psychiatry*, 168(1): 65-72.
- Baldwin, H., Biehal, N., Allgar, V., Cusworth, L., & Pickett, K. (2020). Antenatal risk factors for child maltreatment: linkage of data from a birth cohort study to child welfare records. *Child Abuse & Neglect*, *107*, 104605.
- Chandan, J. S., Thomas, T., Gokhale, K. M., Bandyopadhyay, S., Taylor, J., & Nirantharakumar, K. (2019). The burden of mental ill health associated with childhood maltreatment in the UK, using The Health Improvement Network database: a population-based retrospective cohort study. *The Lancet Psychiatry*, 6(11), 926-934.
- Clark, C, Caldwell, T, Power C, Stansfeld, S. A (2010) Does the influence of childhood adversity on psychopathology persist across the lifecourse? A 45-year prospective epidemiologic study. *Annals of epidemiology*, 20(5): 385-394.
- Collishaw S, Dunn J, O’Connor TG, Golding J (2007) Maternal childhood abuse and offspring adjustment over time. *Development and psychopathology*, 19(02): 367-383.
- Denholm R, Power C, Li L (2013) Adverse childhood experiences and child-to-adult height trajectories in the 1958 British birth cohort. *International journal of epidemiology*, 42(5): 1399-1409.
- Denholm R, Power, C, Thomas C, Li L (2013) Child maltreatment and household dysfunction in a British birth cohort. *Child abuse review*, 22(5): 340-353.
- Fisher HL, Caspi A, Moffitt TE, Wertz J, Gray R, Newbury J, et al. (2015) Measuring adolescents’ exposure to victimization: The Environmental Risk (E-Risk) Longitudinal Twin Study. *Development and psychopathology*, 27(4pt2): 1399-1416.
- Ford E, Clark C, Stansfeld SA (2011) The influence of childhood adversity on social relations and mental health at mid-life. *Journal of affective disorders*, 133(1): 320-327.
- Geoffroy MC, Pereira SP, Li L, Power C (2016) Child neglect and maltreatment and childhood-to-adulthood cognition and mental health in a prospective birth cohort. *Journal of the American Academy of Child Adolescent Psychiatry*, 55(1): 33-40.
- González-Izquierdo, A., Woodman, J., Copley, L., van der Meulen, J., Brandon, M., Hodes, D., … & Gilbert, R. (2010). Variation in recording of child maltreatment in administrative records of hospital admissions for injury in England, 1997–2009. *Archives of disease in childhood*, *95*(11), 918-925.
- Gonzalez-Izquierdo, A., Cortina-Borja, M., Woodman, J., Mok, J., McGhee, J., Taylor, J., … & Gilbert, R. (2014). Maltreatment or violence-related injury in children and adolescents admitted to the NHS: comparison of trends in England and Scotland between 2005 and 2011. *BMJ open*, *4*(4).
- Hanlon, P., McCallum, M., Jani, B. D., McQueenie, R., Lee, D., & Mair, F. S. (2020). Association between childhood maltreatment and the prevalence and complexity of multimorbidity: A cross-sectional analysis of 157,357 UK Biobank participants. *Journal of comorbidity*, *10*, 2235042X10944344.
- Ho, F. K., Celis-Morales, C., Gray, S. R., Petermann-Rocha, F., Lyall, D., Mackay, D., … & Pell, J. P. (2020). Child maltreatment and cardiovascular disease: quantifying mediation pathways using UK Biobank. *BMC medicine*, *18*(1), 1-10.
- Jaffee SR, Caspi A, Moffitt TE, Polo-Tomas M, Price TS, Taylor A (2004) The limits of child effects: evidence for genetically mediated child effects on corporal punishment but not on physical maltreatment. *Developmental psychology*, 40(6): 1047.
- Kelly-Irving M, Lepage B, Dedieu D, Lacey R, Cable N, et al. (2013) Childhood adversity as a risk for cancer: findings from the 1958 British birth cohort study. *BMC Public Health*, 13(1): 1.
- Li L, Denholm, R, Power C (2014) Child maltreatment and household dysfunction: associations with pubertal development in a British birth cohort. *International journal of epidemiology*, dyu071.
- Mc Grath-Lone, L., Dearden, L., Nasim, B., Harron, K., & Gilbert, R. (2016). Changes in first entry to out-of-home care from 1992 to 2012 among children in England. *Child abuse & neglect*, *51*, 163-171.
- Mc Grath-Lone, L., Dearden, L., Harron, K., Nasim, B., & Gilbert, R. (2017). Factors associated with re-entry to out-of-home care among children in England. *Child abuse & neglect*, 63, 73-83.
- Murray, E. T., Lacey, R., Maughan, B., & Sacker, A. (2020a). Association of childhood out-of-home care status with all-cause mortality up to 42-years later: Office of National Statistics Longitudinal Study. *BMC public health*, *20*, 1-10.
- Murray, E. T., Lacey, R., Maughan, B., & Sacker, A. (2020b). Non-parental care in childhood and health up to 30 years later: ONS Longitudinal Study 1971–2011. *European Journal of Public Health*, *30*(6), 1121-1127.
- Newbury, J. B., Arseneault, L., Moffitt, T. E., Caspi, A., Danese, A., Baldwin, J. R., & Fisher, H. L. (2018). Measuring childhood maltreatment to predict early-adult psychopathology: comparison of prospective informant-reports and retrospective self-reports. *Journal of psychiatric research*, *96*, 57-64.
- Power C, Pereira SMP, Li L (2015) Childhood maltreatment and BMI trajectories to mid-adult life: follow-up to age 50y in a British birth cohort. *PloS one*, 10(3): e0119985
- Radford, L., Corral, S., Bradley, C., & Fisher, H. L. (2013). The prevalence and impact of child maltreatment and other types of victimization in the UK: Findings from a population survey of caregivers, children and young people and young adults. *Child abuse & neglect*, *37*(10), 801-813.
- Schurer, S., Trajkovski, K., & Hariharan, T. (2019). Understanding the mechanisms through which adverse childhood experiences affect lifetime economic outcomes. *Labour Economics*, *61*, 1017-43.
- Sidebotham P, Golding J, ALSPAC Study Team (2001) Child maltreatment in the “Children of the Nineties”: A longitudinal study of parental risk factors. *Child Abuse and Neglect,* 25(9): 1177-1200.
- Sidebotham P, Heron J, Golding J, ALSPAC Study Team (2002) Child maltreatment in the “Children of the Nineties:” deprivation, class, and social networks in a UK sample. *Child Abuse and Neglect,* 26(12): 1243-1259.
- Sidebotham P, Heron J, Teamc, T. A. S (2003) Child maltreatment in the “children of the nineties:” the role of the child. *Child Abuse and Neglect,* 27(3): 337-352.
- Sidebotham P, Heron J, ALSPAC Study Team (2006) Child maltreatment in the “children of the nineties”: A cohort study of risk factors. *Child Abuse and Neglect,* 30(5): 497-522.
- Soares, A. L. G., Hammerton, G., Howe, L. D., Rich-Edwards, J., Halligan, S., & Fraser, A. (2020). Sex differences in the association between childhood maltreatment and cardiovascular disease in the UK Biobank. *Heart*, *106*(17), 1310-1316.
- Solís CB, Kelly-Irving M, Fantin R, Darnaudéry M, Torrisani J, et al. (2015) Adverse childhood experiences and physiological wear-and-tear in midlife: Findings from the 1958 British birth cohort. *Proceedings of the National Academy of Sciences*, 112(7): E738-E746.
- Suderman M, Borghol N, Pappas JJ, Pereira SMP, Pembrey M, et al. (2014) Childhood abuse is associated with methylation of multiple loci in adult DNA. *BMC medical genomics*, 7(1): 1.
- Thomas C, Hyppönen E, Power C (2008) Obesity and type 2 diabetes risk in midadult life: the role of childhood adversity. *Pediatrics*, 121(5): e1240-e1249.
- Woodman J, Freemantle N, Allister J, de Lusignan S, Gilbert R, Petersen I (2012) Variation in recorded child maltreatment concerns in UK primary care records: a cohort study using The Health Improvement Network (THIN) database. *PLoS One*, 7(11): e49808.

#### Economics Studies of Child Maltreatment

- Bald, A., Chyn, E., Hastings, J. S., & Machelett, M. (2019). *The causal impact of removing children from abusive and neglectful homes* (No. w25419). National Bureau of Economic Research.
- Bitler MP, Zavodny M (2004) Child maltreatment, abortion availability, and economic conditions. *Review of Economics of the Household*, 2(2): 119-141.
- Currie J, Tekin E (2012) Understanding the cycle childhood maltreatment and future crime. *Journal of Human Resources*, 47(2): 509-549.
- Doyle Jr, J. J. (2007). Child protection and child outcomes: Measuring the effects of foster care. *American Economic Review*, *97*(5), 1583-1610.
- Doyle Jr, J. J. (2008). Child protection and adult crime: Using investigator assignment to estimate causal effects of foster care. *Journal of Political Economy*, *116*(4), 746-770.
- Doyle Jr, J. J., & Aizer, A. (2018). Economics of child protection: Maltreatment, foster care, and intimate partner violence. *Annual review of economics*, *10*, 87-108.
- Fletcher JM (2009) Childhood mistreatment and adolescent and young adult depression. *Social Science Medicine*, 68(5): 799-806.
- Fletcher JM, Schurer S (2015) Childhood Origins of Adulthood Noncognitive Skills: The Role of Chronic Health Problems and Exposure to Maltreatment (ARC lifecourse centre dp No. 2015-23).
- Gross M, Baron EJ (2021) Temporary stays and persistent gains: The causal effects of foster care. *American Economic Journal: Applied*, forthcoming.
- Paxon C, Waldfogel J (1999) Parental resources and child abuse and neglect. *The American Economic Review*, 89(2): 239-244.
- Paxon C, Waldfogel J (2002) Work, welfare, and child maltreatment. *Journal of Labor Economics*, 20(3): 435-474.
- Paxon C, Waldfogel J (2003) Welfare reforms, family resources, and child maltreatment. *Journal of Policy Analysis and Management*, 22(1): 85-113.
- Reeve R, Gool K (2013) Modelling the Relationship between Child Abuse and Long-Term Health Care Costs and Wellbeing: Results from an Australian Community-Based Survey. *Economic Record*, 89(286): 300-318.
- Schurer, S., Trajkovski, K., & Hariharan, T. (2019). Understanding the mechanisms through which adverse childhood experiences affect lifetime economic outcomes. *Labour Economics*, *61*, 1017-43.

#### Cost Studies

- Alabama Children’s Trust Fund, et al. (2007) *The costs of child abuse vs. child abuse prevention: Alabama’s experience*. Alabama: University of Alabama, Center for Business and Economic Research.
- Corso PS, Lutzker JR (2006) The need for economic analysis in research on child maltreatment. *Child Abuse and Neglect*, 30(7): 727-83.
- Corso PS, Mercy JA, Simon TR, Finkelstein EA, Miller TR (2007) Medical costs and productivity losses due to interpersonal and self-directed violence in the United States. *American Journal of Preventive Medicine,* 32(6): 474-48.
- Corso PS, Fertig AR (2010) The economic impact of child maltreatment in the United States: are the estimates credible?. *Child Abuse and Neglect,* 34(5): 296-304.
- Dolezal T, McCollum D, Callahan M (2009) *Hidden costs in health care: the economic impact of violence and abuse*. Eden Prairie, Minnesota: Academy of Violence and Abuse.
- Dubowitz, H (1990) Costs and effectiveness of interventions in child maltreatment. *Child Abuse and Neglect*, 14(2): 177-86.
- Fang X, Brown DS, Florence CS, Mercy JA (2012) The economic burden of child maltreatment in the United Stated and implications for prevention. *Child Abuse and Neglect,* 36(2):156-65.
- Fang X, Fry DA, Brown DS, Mercy JA, Dunne MP, et al. (2015) The burden of child maltreatment in the East Asia and Pacific region. *Child Abuse and Neglect*, 42: 146-62.
- Fang X, Fry DA, Ji K, Finkelhor D, Chen J, et al. (2015) The burden of child maltreatment in China: a systematic review. *Bulletin of the World Health Organization*, 93(3): 176-185 C.
- Florence C, Brown DS, Fang X, Thompson HF (2013) Health care costs associated with child maltreatment: impact on Medicaid. *Pediatrics,* 132(2): 312-318.
- Giardino AP, Montoya LA, Richardson AC, Leventhal JM (1999) Funding realities: child abuse diagnostic evaluations in the health care setting. *Child Abuse and Neglect*, 23(6): 531-38.
- Habetha, S., Bleich, S., Weidenhammer, J., & Fegert, J. M. (2012). A prevalence-based approach to societal costs occurring in consequence of child abuse and neglect. *Child and adolescent psychiatry and mental health*, *6*(1), 35.
- Hennes H, Kini N, Palusci VJ (2001) The epidemiology, clinical characteristics and public health implications of shaken baby syndrome*. Journal of Aggression, Maltreatment and Trauma*, 5(1), 19-40.
- Irazuzta JE, McJunkin JE, Danadian K, Arnold F, Zhang J (1997) Outcome and cost of child abuse. *Child Abuse and Neglect*, 21(8): 751-7.
- Kahui S, Suzanne S (2014) *Measuring the Economic Costs of Child Abuse and Intimate Partner Violence to New Zealand*. New Zealand: The Glenn Inquiry.
- McCarthy, M. M., Taylor, P., Norman, R. E., Pezzullo, L., Tucci, J., & Goddard, C. (2016). The lifetime economic and social costs of child maltreatment in Australia. *Children and youth services review*, 71, 217-226.
- NICE, (2009) *Costing statement: when to suspect child maltreatment*. London: RCOG Press. [Online] Available from: https://www.nice.org.uk/guidance/cg89/evidence/full-guideline-pdf-243694625
- Perryman Report (2014) *Suffer the Little Children: An assessment of the Economic Cost of Child Maltreatment.* Texas: The Perryman Group.
- Peterson C, Xu L, Florence C, Parks SE, Miller TR, et al. (2014) The medical cost of abusive head trauma in the United States. *Pediatrics*, 134(1): 91-99.
- Peterson C, Xu L, Florence C, Parks SE (2015) Annual Cost of U.S. Hospital Visits for Pediatric Abusive Head Trauma. *Child Maltreatment*, 20(3): 162-9.
- Peterson C, Florence, C, Klevens, J (2018) The economic burden of child maltreatment in the United States, 2015. *Child Abuse and Neglect*, 86: 178–183.
- Saied-Tessier A (2014) *Estimating the costs of child sexual abuse in the UK*. London: NSPCC.
- Sethi, D., Bellis, M., Hughes, K., Gilbert, R., Mitis, F., & Galea, G. (2013). *European report on preventing child maltreatment*. World Health Organization. Regional Office for Europe.
- Summers CL, Molyneux EM (1992) Suspected child abuse: cost in medical time and finance. *Archives of disease in childhood,* 67(7): 905-10.
- Wada I, Ataru I (2014) The social costs of child abuse in Japan. *Children and Youth Services Review*, 46 (2014): 72-77.
- Wang, C., Holton, J (2007) *Total estimated cost of child abuse and neglect in the United States*. Chicago: Prevent Child Abuse America.
- Wekerle C (2011) The dollars and senselessness in failing to prioritize childhood maltreatment prevention. *Child Abuse and Neglect,* 35(3): 159-61.
- World Health Organization, Violence and Injury Prevention Programme (2007) *Preventing child maltreatment in Europe: a public health approach: policy briefing*. Copenhagen: World Health Organization Regional Office for Europe.
- Woodman J (2011) Healthcare use by children fatally or seriously harmed by child maltreatment: analysis of a national case series 2005-2007. *Archives of Disease in Childhood*. 96(3): 270-5.

### Appendix 2: Definition and construction of the child maltreatment measures

#### National Child Development Study

##### Neglect

An individual is defined as having been neglected in childhood if he/she reports that *any* of the three following conditions is true, as compared to none of them being true.^1^

1. “Mother (or mother figure) not at all affectionate towards me up to age 16” [Variable CHAD4].
2. “Father (or father figure) not at all affectionate towards me up to age 16” [Variable CHAD1].
3. “I was neglected up to age 16” [Variable CHAD9C].

**Table 2A:** Mother Neglect in NCDS.

| **Table 2A: Mother Neglect in NCDS** |  |  |
| --- | --- | --- |
| Father (figure) was affectionate towards me |  |  |
|  | Freq. | Percent |
| A lot | 5,131 | 55.11% |
| Somewhat | 2,437 | 26.2% |
| A little | 1,287 | 13.8% |
| Not at all | 327 | 3.5% |
| I had no father figure<sup>2</sup> | 59 | 0.6% |
| Can't say | 70 | 0.8% |
| Total | 9,311 | 100.0% |

**Table 2B:** Father Neglect in NCDS.

| **Table 2B: Father Neglect in NCDS** |  |  |
| --- | --- | --- |
| Mother (figure) was affectionate towards me |  |  |
|  | Freq. | Percent |
| A lot | 3,081 | 33.1% |
| Somewhat | 2,405 | 25.8% |
| A little | 2,563 | 27.5% |
| Not at all | 851 | 9.1% |
| I had no mother figure | 253 | 2.7% |
| Can't say | 158 | 1.7% |
| Total | 9,311 | 100.0% |

**Table 2C:** Neglect in NCDS.

| **Table 2C: Neglect in NCDS** |  |  |
| --- | --- | --- |
| I was neglected |  |  |
|  | Freq. | Percent |
| Yes | 230 | 2.5% |
| No | 8,859 | 95.2% |
| Can't say | 220 | 2.4% |
| Total | 9,309 | 100.0% |

**Table 2D:** Any neglect in NCDS.

| **Table 2D: Any neglect in NCDS** |  |  |
| --- | --- | --- |
|  | Freq. | Percent |
| Yes | 1,154 | 12.4% |
| No | 7,950 | 85.4% |
| Can't say | 204 | 2.2% |
| Total | 9,308 | 100.0% |

<sup>1</sup> A separate variable has been included as control in all the estimated models in case the answers include both “I had no mother/father figure” and “can't say”.
<sup>2</sup> We kept categories ‘No father/mother figure’ and ‘Can't say’ separately from affirmative or negative answers to CM questions. In other words, for each CM measure, in the regression analyses the baseline category was not suffering CM, compared each of categories 1) ‘suffering CM’; 2) ‘No father/mother figure’; 3) ‘Can't say’. In the paper we present results for (1); results for (2) and (3) are available upon request.

##### Emotional Abuse

An individual is defined as having been emotionally abused in childhood if he/she reports that *any* of the three following conditions is true, as compared to none of them being true. ^3^

1. “I was verbally abused by a parent (or parent figure) up to age 16” [Variable CHAD9F].
2. “I suffered humiliation, ridicule, bullying or mental cruelty from a parent (or parent figure) up to age 16” [CHAD9G].
3. “I witnessed physical or sexual abuse of others in my family up to age 16” [CHAD9H].

**Table 2E:** Verbal abuse in NCDS.

| **Table 2E: Verbal abuse in NCDS** |  |  |
| --- | --- | --- |
| I was verbally abused by a parent (figure) |  |  |
|  | Freq. | Percent |
| Yes | 746 | 8.0% |
| No | 8,388 | 90.1% |
| Can't say | 175 | 1.9% |
| Total | 9,309 | 100.0% |

**Table 2F:**
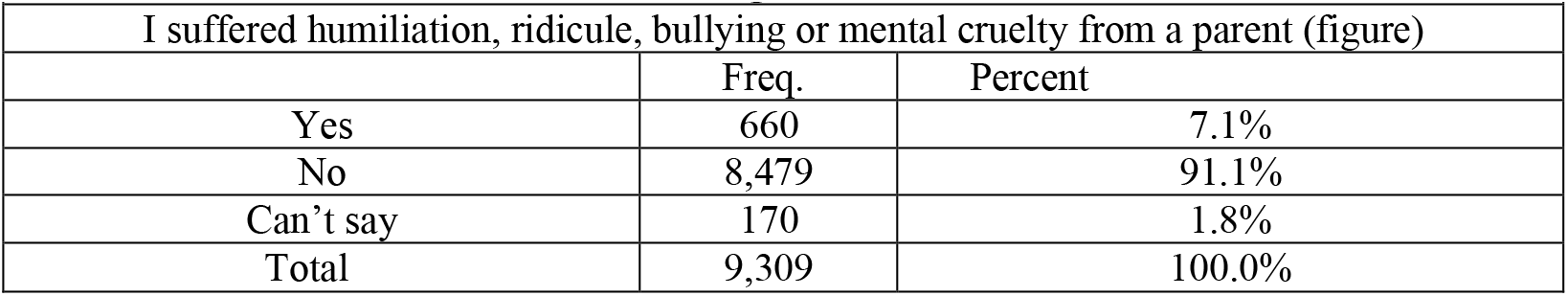
Suffering humiliation in NCDS.

**Table 2G:** Witnessed abuse in NCDS.

| **Table 2G: Witnessed abuse in NCDS** |  |  |
| --- | --- | --- |
| I witnessed physical or sexual abuse of others in my family |  |  |
|  | Freq. | Percent |
| Yes | 559 | 6.0% |
| No | 8,683 | 93.3% |
| Can't say | 67 | 0.7% |
| Total | 9,309 | 100.0% |

**Table 2H:** Any emotional abuse in NCDS.

| **Table 2H: Any emotional abuse in NCDS** |  |  |
| --- | --- | --- |
|  | Freq. | Percent |
| Yes | 1,154 | 12.4% |
| No | 7,950 | 85.4% |
| Can't say | 204 | 2.2% |
| Total | 9,308 | 100.0% |

##### Physical Abuse

An individual is defined as having been physically abused in childhood if he/she reports yes to “I was physically abused by a parent – punched, kicked or hit or beaten with an object, or needed medical treatment up to age 16” [Variable CHAD9I].^4^

**Table 2I:** Physical abuse in NCDS.

| **Table 2I: Physical abuse in NCDS** |  |  |
| --- | --- | --- |
| I was physically abused by a parent (figure) |  |  |
|  | Freq. | Percent |
| Yes | 559 | 6.0% |
| No | 8,663 | 93.1% |
| Can't say | 87 | 0.9% |
| Total | 9,309 | 100.0% |

<sup>3</sup> A separate variable has been included as control in all the estimated models in case the answers include both “no” and “can’t say”.
<sup>4</sup> A separate variable has been included as control in all the estimated models in case the answer is “can’t say”.

##### Sexual Abuse

An individual is defined as having been sexually abused in childhood if he/she reports yes to “I was sexually abused by a parent (or parent-figure) up to age 16” [Variable CHAD9K].

**Table 2J:** Sexual abuse in NCDS.

| **Table 2J: Sexual abuse in NCDS** |  |  |
| --- | --- | --- |
| I was sexually abused by a parent (figure) |  |  |
|  | Freq. | Percent |
| Yes | 143 | 1.5% |
| No | 9,124 | 98.0% |
| Can't say | 42 | 0.5% |
| Total | 9,309 | 100.0% |

##### Global Measure of Abuse

An individual is defined as having been abused or neglected in childhood if he/she reports suffering any form of abuse or neglect up to age 16.

**Table 2K:** Global abuse in NCDS.

| **Table 2K: Global abuse in NCDS** |  |  |
| --- | --- | --- |
| I was abused by a parent (figure) |  |  |
|  | Freq. | Percent |
| Yes | 1,914 | 20.6% |
| No | 7,085 | 76.1% |
| Can't say | 308 | 3.3% |
| Total | 9,307 | 100.0% |

#### English Longitudinal Study of Ageing

The ELSA includes retrospective measures of CM (physical abuse and neglect only), which were asked in the life history module in wave 3. The following definitions were used.

<u>Neglect</u>: an individual is defined as having been neglected in childhood if he/she answers “agree or strongly agree” to the question “Mother (mother figure) or Father (father figure) seemed emotionally cold to me”.

<u>Physical Abuse</u>: an individual is defined as having been physically abused in childhood if he/she answers yes to the question “When you were aged under 16, were you physically abused by your parents”.

**Table 2L:**
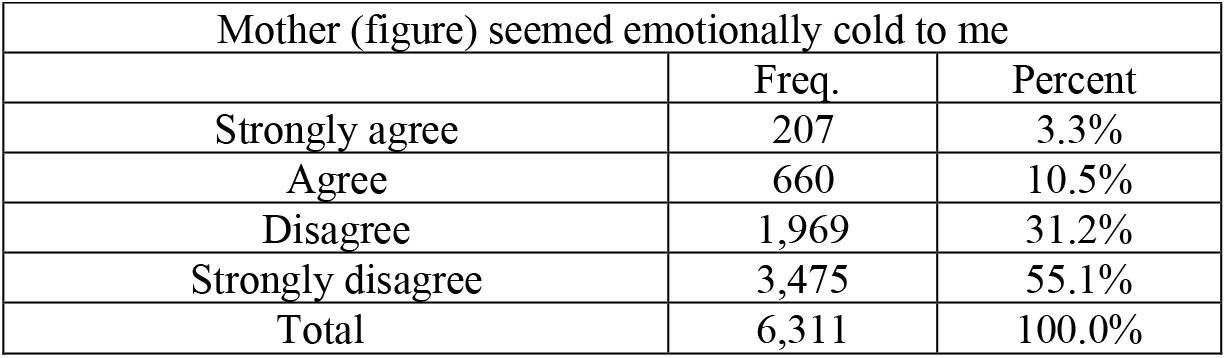
Mother Neglect in ELSA.

| **Table 2L: Mother Neglect in ELSA** |  |  |
| --- | --- | --- |
| Mother (figure) seemed emotionally cold to me |  |  |
|  | Freq. | Percent |
| Strongly agree | 207 | 3.3% |
| Agree | 660 | 10.5% |
| Disagree | 1,969 | 31.2% |
| Strongly disagree | 3,475 | 55.1% |
| Total | 6,311 | 100.0% |

**Table 2M:** Father Neglect in ELSA.

| **Table 2M: Father Neglect in ELSA** |  |  |
| --- | --- | --- |
| Father (figure) seemed emotionally cold to me |  |  |
|  | Freq. | Percent |
| Strongly agree | 236 | 3.9% |
| Agree | 743 | 12.1% |
| Disagree | 2,296 | 37.4% |
| Strongly disagree | 2,862 | 46.7% |
| Total | 6,137 | 100.0% |

**Table 2N:**
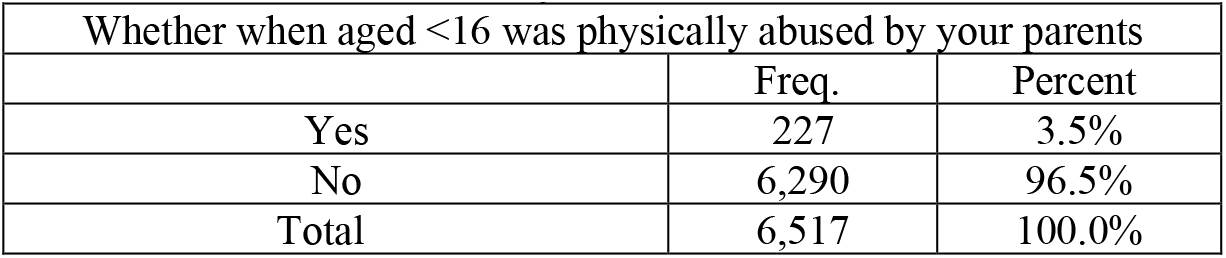
Physical Abuse in ELSA.

### Appendix 3: Detailed description of methods used to estimate costs

#### Costs included in the analysis

We assessed the lifetime cost of child maltreatment in the UK adopting a societal perspective, including costs to the health care, social care, education and criminal justice sectors, and to the wider economy in terms of lost productivity. Based on the literature review and analyses described above we included: short-term health-related costs; long-term health-related costs; criminal justice costs; children’ social care system costs; special education costs; and, productivity losses due to reduced employment.

Short-term health-related costs include costs associated with unplanned hospital admissions for injuries, and health care and criminal justice costs associated with mental health disorders arising from child maltreatment.

Long-term health related costs include costs due to depression, anxiety, smoking and alcohol abuse, which were found to be related to child maltreatment (the global measure) in the econometric analysis. We did not include costs associated with obesity, hypertension, type 2 diabetes, raised cholesterol, cancer and height as these were not significantly related to the global measure of child maltreatment. We have included costs of other conditions that are related to depression, anxiety, smoking and alcohol abuse where these are included in published estimates (e.g., the cost of developing lung cancer among smokers is included in the smoking-related costs).

Productivity losses might arise from child maltreatment due to premature mortality, days off work, reduced employment and reduced wages among those who are employed. In our analysis productivity losses were captured in two ways. First, in terms of reduced employment. This was based on the findings of the econometric analysis. Second, we included productivity losses arising from days off work and premature mortality due to long-term health-related problems associated with child maltreatment. We were not able to evaluate days off work among those who are employed arising from child maltreatment directly, as these data were not available in the datasets we used for our analysis, but we did include these costs in our calculation of long-term health-related costs where they were included in published cost estimates (e.g., absenteeism due to depression). To avoid double counting, when we included productivity losses from long-term health-related problems we removed the productivity losses due to reduced unemployment. We found that child maltreatment was not associated with lower wages among those who were employed, so we did not include this effect.

We did not include costs associated with drug use as we did not have data on this in our econometric analysis. We did not evaluate the impact of child maltreatment on education attainment, but we included special education costs associated with child maltreatment and accounted for education effects indirectly in our calculation of productivity losses. We did not evaluate the impact of child maltreatment on the likelihood and cost of divorce as it was unclear how we would value these costs. Our econometric analysis showed that child maltreatment is associated with increased disability throughout the life course and the receipt of disability benefits. Health-related costs are included in our analysis but we did not include disability benefits as these are transfer payments^5^ (Hogson and Meiners, 1982’ Luce et al 1996, Choi et al 1997).

#### Costing methodology

We used the econometric analysis described above, supplemented with the limited but best available published evidence to develop our cost estimates. We used the findings from the analysis of the global measure of child maltreatment, and preferred the NCDS results over the ELSA results as the former included more types of child maltreatment. The only exception to this is that we used the ELSA results for heavy drinking as this was felt to be a more appropriate measure of alcohol abuse than the NCDS measure. We used the coefficients from the most detailed econometric model (specification (4) of the NCDS analysis; (2) of the ELSA analysis). While some relevant variables were available in the datasets we analysed to measure the short-terms costs described above the data were not sufficiently detailed (e.g., in terms of the timing of child maltreatment). Therefore we based our calculations of short-term costs on published figures. Secondary evidence was identified via searches of several databases including PubMed, the NHS Economic Evaluations Database, EconLit, Google Scholar and Google. For each cost category we used published estimates from previous cost of illness studies where possible. We used data that were specific to the UK as opposed to countries outside of the UK because health care, social care, education and criminal justice systems vary between countries and costs in other countries are unlikely to apply to the UK. The only exception to this was that in sensitivity analyses we applied costs of child maltreatment calculated for the USA by Fang and colleagues (Fang et al, 2012) to the UK to compare differences between the two studies. Costs vary over time and we used recent data whenever possible, though as noted below several of the data sources are dated.

We did not evaluate costs by type and severity of child maltreatment and instead opted for an overall maltreatment estimate. One reason is that many of the studies used to provide inputs into the calculations did not distinguish by type of maltreatment, so disaggregating costs would be difficult. In our econometric analysis we were able to analyse the impact of different types of maltreatment on long-term health outcomes and labour market outcomes, but small numbers of cases for some types of maltreatment meant the analysis was underpowered. Another issue is that there may be overlap between different types of maltreatment making it difficult to attribute costs to individual types of maltreatment.

For several of the cost components we required longitudinal data on the number of events for new cases of child maltreatment in a single year to be able to accurately calculate incidence-based costs. These data were not available so as noted below we were required to make ‘steady-state’ assumptions, as have been used by other researchers. Using this approach the number of events in a single year across all victims of child maltreatment is a proxy for all events over time among new victims in that year. This assumption requires that the number of events remains fairly constant over time, which may be problematic for some of the cost components considered here.

Our aim was to estimate the lifetime cost per victim of child maltreatment, for several of the cost components data were only available at the aggregate level across all victims. Therefore to calculate costs per victim we were required to divide these costs through by an estimate of the number of new cases of child maltreatment. This number is uncertain.

All costs were estimated in UK£ and adjusted to the 2015 prices (our reference year) where necessary using gross domestic product deflators^6^.

We assumed that when children are maltreated the average age at which it starts is 6 years (Department for Education, 2016)^7^. We used this as the starting point to calculate lifetime costs per victim, but acknowledge there is some uncertainty with this figure. There is no data source that tells at what age abuse typically starts. Our assumption that it starts on average at 6 years of age is based on published child protection statistics that are available in each of the four nations of the UK and reflects the age children become subject to a child protection plan or come on to the child protection register. We have looked at data for England, Wales Northern Ireland and Scotland and taken weighted averages. Where age ranges were available mid-points were used to calculate weighted averages. This assumption is conservative because we know the age of the child at the end of the year as opposed to when the child become subject to a protection plan. Also the abuse will have started before children become subject to a protection plan. The abuse could have just started, or it could have been happening for a number of years.

Costs are presented in present value terms; future costs up to 30 years from the age of 6 are discounted using an annual rate of 3.5% (NICE, 2013)^8^; for costs incurred between 31 and 74 years from age 6 we applied an annual discount rate of 3%; for those over 75 years we applied an annual rate of 2.5%.^9^ As noted above, and described in more detail below, there is uncertainty in our estimates, reflecting data limitations. We therefore provide central estimates and undertook extensive sensitivity analysis to take this into account.

### Lifetime costs per victim of non-fatal child maltreatment

#### Short-term health-related costs

Short-term health-related costs of child maltreatment refer to health care and associated costs up to the age of 18. We include costs incurred by treating injuries from maltreatment and to treat mental health problems associated with maltreatment (Summers and Molyneaux, 1992; Hernes, 2001, Corso and Fertig, 2010; Brown, Fang et al, 2011; Fan, Brown et al, 2012, Florence, Brown et al, 2013; Peterson, Xu et al, 2014; Saied-Tessier 2014, Boschung, 2015)

#### Unplanned hospital admissions for injuries

We valued short-term health care costs in terms of unplanned hospital admissions for maltreatment or violence-related injuries. According to Gonzales-Izquierdo et al (Gonzales-Izquierdo, Cortina Borja et al, 2014) between 1 January 2005 and 31 March 2012 there were 61,574 unplanned hospital admissions (ordinary admissions and day cases) for maltreatment or violence-related (MVR) injury in children aged 18 years or younger in England and Scotland, where MVR diagnoses were defined using previously published and validated methods.

We applied the unplanned admission rate for maltreatment or violence-related injuries for England in 2011 by age groups reported by Gonzales-Izquierdo et al to mid-year UK population estimates and estimated there were 8,685 unplanned admissions in 2015 in the UK. Ideally we would have longitudinal data on the number admissions for new cases of child maltreatment in a single year. These data are not available so we use a ‘steady-state’ methodology also used by other researchers to estimate the lifetime costs of a disease or problem when longitudinal data are not available (Barnett, Birnbaum et al, 2000, Leong et al, 2003, Fang, Brown et al, 2012). Making this assumption, the total admissions in one year for all victims is a proxy for all admissions over time for new victims in that year. We therefore assume in the absence of data that these admissions relate to new cases of child maltreatment. The national average unit cost of non-elective long stay and non-elective short stay admissions for paediatric injuries was £853 (Department of Health, 2015). Hence the cost of unplanned hospital admissions for maltreatment or violence-related injuries among new cases of maltreatment in 2015 is estimated to be 8,685*£853 = £7,405,733. This figure was divided by an estimate of the incidence of child maltreatment based on the number of children coming onto child protection plans or registers in 2015 minus the number of re-registrations (61,894 (Department of Education, 2016)) to give an estimate of the average cost per victim of £120.

Note this calculation assumes that admissions rates for maltreatment or violence-related injuries across the whole of the UK are the same as for England, which may not be true. Also, this figure is an underestimate of the costs of treating child-maltreatment-related injuries as it only includes hospital costs; we did not include other health care costs, e.g., from primary care contacts and outpatient attendances, as we were unable to attribute these to child maltreatment. We explore the impact of including these costs in sensitivity analyses.

#### Short-term mental health problems

Maltreatment is commonly associated with mental health-related responses to stress and trauma, including depression, anxiety, post-traumatic stress disorder and behavioural disorders, but UK evidence is sparse on the incremental effect of child maltreatment on mental health problems in children and young people.

To assess the incremental effect of child maltreatment on mental health problems we used the prevalence of mental health problems in children in care minus the prevalence of those not in care (Meltzer, Gatward et al, 2003). This assumes the prevalence of mental health problems for children who have been maltreated is the same as those in care. In a study of mental health problems, service use and costs among children in foster care, 93% of the sample had suffered some form of abuse or neglect (Minnis, Everett et al, 2006). This approach also assumes that if those who were maltreated were not maltreated then the prevalence in this group would be the same as those not in care. Using this approach, the incremental probability of suffering emotional disorders, conduct disorders and hyperkinetic disorders is summarised in Table A2 by age group. In the absence of data we assumed the effect in children aged 11-15 years applies to those up to the age of 18.

We combined these figures with costs reported by Snell and colleagues (Snell, Knapp et al., 2013). Mean annual costs of hyperkinetic disorders, conduct disorders and emotional disorders in 2015 prices were £3,605, £2,153 and £1,351, respectively, including primary care, paediatric health services and mental services costs.

We then estimated the incremental cost of child maltreatment on mental health problems in children and young people by multiplying the estimated incremental effect of child maltreatment on the probability of each disorder by the annual costs for each year from 6 to 18 years. Summing across the different types of disorder the total discounted costs were £11,453 per victim up to age 18 (i.e., over 12 years).

There is evidence that maltreatment may lead to increased delinquency and crime(Rutter, 1998), though data for the UK are limited. We estimated criminal justice system costs attributable to mental health problems in children and young people based on mean criminal justice system costs per year associated with conduct disorder reported by Scott and colleagues (Scott, Knapp et al, 2001), (£3,436 in 2015 prices). We multiplied this figure by the increased probability of conduct disorder associated with child maltreatment in Table A5. Applying these figures from ages 6 to 18, the discounted cost was £7,100 per victim (i.e., over 12 years).

The total cost of mental health problems in maltreated children is the sum of the health service and criminal justice system costs, or £18,553 per victim. The main uncertainty in this analysis is the assumption concerning the impact of child maltreatment on mental health problems. We explore this further in sensitivity analysis. There is also evidence that mental health problems in childhood are associated with significant societal costs in adulthood, but we included some of these costs in the long-term health-related costs below.

#### Long term health-related costs

We considered the long-term impact of child maltreatment on anxiety, depression, smoking and alcohol abuse in adulthood based on the findings of the econometric analysis described in the previous section.

#### Anxiety

McCrone and colleagues (McCrone, 2008) calculated that the average cost of anxiety per year per person with anxiety who is in contact with the health service is £1,282 (2015 prices). This includes direct health and social care costs, informal care costs and criminal justice system costs. McCrone and colleagues also report productivity losses due to anxiety, but these are attributed to the impact of anxiety on the probability of employment and so we do not include them to avoid double counting. Fineberg and colleagues (Fineberg, Haddad et al, 2013) calculated the total societal cost of anxiety per patient and reported that 46.4% of the total was due to productivity losses arising from work absence and early retirement; from this study we calculated that the productivity losses due to anxiety are £661 per person (2015 prices).

The results of the NCDS analysis using the global CM measure show that child maltreatment was positively correlated with anxiety at ages 33, 42, 50 and 55. There was a non-significant effect at age 23 (appendix, Table A2); we therefore assumed the marginal effect of child maltreatment on the probability of anxiety was zero at this age. The marginal effect of child maltreatment at each year of age over the lifetime was calculated by assuming a straight line relation between the marginal effects at ages 23, 33, 42, 50 and 55. We assumed the effect of child maltreatment before the age of 23 was captured by the short-term health-related costs described above and so did not include an impact here. We assumed the marginal effect at age 55 was constant over the remaining lifetime.

The mean incremental lifetime cost of anxiety in maltreated children was calculated as the product of the average costs per year (health care costs and productivity losses) and the marginal effect of child maltreatment on the probability of anxiety at each year of age. Due to retirement, after age 67 we did not include productivity losses.

Using this approach, the average discounted lifetime incremental cost of anxiety per victim of child maltreatment was estimated to be £954.

#### Depression

To estimate the costs of depression associated with child maltreatment we used the same approach as for anxiety, described above. McCrone and colleagues (McCrone, 2008) calculated that the cost of depression per year per person with depression who is in contact with the health service was £2,418 (2015 prices). The cost of depression includes direct health and social care costs, informal care costs and criminal justice system costs. Productivity losses due to depression were taken from Thomas and colleagues (Thomas, 2000) who calculated that costs of lost productivity with depression due to days off work and premature mortality were £4,479 per person with depression per year (2015 prices).

The NCDS analysis showed that child maltreatment was positively correlated with depression at ages 33, 42, 50 and 55, but not at age 23 (Table A2), and we applied the same methodology to calculate the marginal effect of child maltreatment on depression at each year of age.

The discounted lifetime incremental cost of depression per victim of child maltreatment was calculated to be £5,145.

#### Smoking

Action on Smoking and Health (ASH) (Recknoer, 2016)^10^ recently estimated annual costs to society due to smoking. These included the cost to the NHS and social care services of smoking-related health problems, plus costs to society due to smoking-related fires, passive smoking and productivity losses due to smoking breaks, days off sick and early deaths. The total cost in England in 2015 was estimated to be £13.9 billion. The authors also reported an estimated smoking population in England of 7,687,769, indicating a mean societal cost per smoker per year of £1,805 (2015 prices).

Our NCDS analysis showed that child maltreatment was positively correlated with being a current smoker at ages 33, 42 and 50 but not at ages 23 (Table A2) and 55. We assumed the marginal effect of child maltreatment on smoking was zero at ages 23 and 55. The effect of child maltreatment over the lifetime was calculated by assuming a straight line relation between marginal effects at ages 23, 33, 42, 50 and 55. We assumed the effect of child maltreatment on smoking before the age of 23 was zero and that the marginal effect of zero at age 55 was constant for the remaining lifetime.

The mean incremental lifetime cost of smoking in maltreated children was calculated by multiplying the cost per smoker per year by the marginal effect of child maltreatment at each year of age. These were then discounted and summed across the lifetime, yielding a discounted lifetime incremental cost of smoking per victim of child maltreatment of £528.

#### Alcohol abuse

The Cabinet Office (2003) calculated the cost of alcohol misuse to society in England at £11.9 billion per annum (2001 prices), and reported there were 9,114,371 alcohol misusers in the same time period. The costs included NHS costs, lost productivity and crime-related costs. The lost productivity costs included the impact of reduced employment. We removed this element from the total to avoid double counting. We then inflated the figures to 2015 prices and calculated the average cost per alcohol misuser per year to be £2,563.

According to our ELSA analysis, the impact of child maltreatment on the probability of heavy drinking (consuming 2 or more alcoholic drinks a day) was positive and statistically significant. The dataset comprised survey participants aged 50 years or more and we therefore assumed the effect of child maltreatment on alcohol misuse starts at age 50 and remains constant over the remaining lifetime. We assumed the productivity losses apply only up to an assumed retirement age of 67^11^.

We assumed the measure of heavy drinking in the ELSA data was commensurate with the definition of alcohol misuse used to generate the cost estimates and calculated the lifetime cost as the product of the cost per year of alcohol misuse and the marginal effect of child maltreatment on heavy drinking. Future costs were discounted and summed across the lifetime. On this basis the average discounted lifetime incremental cost of alcohol abuse per victim of child maltreatment was estimated to be £537. This figure is probably an underestimate as it assumes there is no impact of child maltreatment on problem drinking before 50 years of age.

#### Criminal justice system costs

Criminal justice system costs included in this category relate to the costs for police, court and penal services for the perpetrators of child abuse. As indicated, criminal justice system costs associated with the consequences of child maltreatment among victims is included in other cost categories. We based our estimates on sanction figures for child sexual abuse, because criminal justice data for other forms of maltreatment were not available. Not all cases of abuse will result in formal criminal proceedings or convictions, therefore proportions were calculated as the ratio of the number of proceeding and convictions to the total number of offences.

In 2013 police recorded 23,300 cases of sexual offences against children in the UK (Saied-Tessier, 2014). In 2013 (the most recent year data are available) there were 4,100 court proceedings and 2,500 convictions registered for sexual offences against children (Saied-Tessier, 2014). In the previous year 1,034 people sentenced for child sexual abuse were admitted onto sexual offender programmes. We calculated the proportion of sexual offence cases that resulted in court proceedings and convictions and applied these proportions to the total number of criminal cases for sexual offences, abuse and neglect against children in 2015. We also applied the proportion of sexual offence cases that led to admission onto sexual offenders programmes in 2013 to the number of criminal cases of sexual offences against children in 2015. The resulting figures, shown in Table 3A, were multiplied by the unit costs of court proceedings, convictions and sex offender treatment programmes (shown in Table 3A in 2015 prices) and summed to calculate total criminal justice system costs. Note the proportion of offences resulting in referral to sex offenders programmes is applied only to sexual abuse cases.

Note that as only a limited number of individuals can be accommodated on sexual offenders programmes the costs reported here are likely to underestimate the financial input needed.

Similar to our treatment of the injury costs above, we make a steady-state assumption and assume the new offences apply to new cases of child maltreatment. Using this assumption, the total number of new offences in one year is used as a proxy for the lifetime number of offences relating to new cases of child maltreatment in that years. This approach also assumes, in the absence of data, that the proportion of prosecuted cases for all type of maltreatment are the same as for sexual offences.

This analysis includes criminal justice costs arising from non-fatal child maltreatment cases. Costs associated with fatal child maltreatment are considered separately below.

Using this approach, the criminal justice system costs due to new cases of child maltreatment in 2015 are estimated to be £267,148,144. This figure was divided by an estimate of the incidence of child maltreatment in 2015 (61,894 (Department for Education (DfE) 2016)) to give an estimate of the average cost per victim of £4,316.

**Table 3A:** Criminal justice system costs due to new cases of child maltreatment in 2015.

|  |  |  |  |
| --- | --- | --- | --- |
|  | **Number** | **Unit cost (£)** | **Total cost (£)** |
| Court proceedings | 10030 | 11,746 | 117,819,570 |
| Convictions | 6116 | 20,362 | 124,535,565 |
| Sex offenders treatment programmes | 2249 | 11,025 | 24,7923,010 |
| Total criminal justice cost |  |  | 267,148,144 |

#### Child social care costs

Child social care costs include the cost of being on a child protection plan or register (initial referral and assessment once maltreatment has been identified, plus ongoing support), and the costs of foster care and local authority residential home care among those who receive it. According to recent data 61% of looked after children in England and 66% in Wales are looked after due to maltreatment. (NSPCC, 2016).

We multiplied the number of children coming onto child protection plans or registers in 2015 minus the number of re-registrations (61,894 (Department of Education, 2016)) by the fixed costs and ongoing support costs obtained from published sources (inflated to 2015 prices; Table 3B). For the ongoing support costs we assumed that all children would receive support for at least 6 months and a proportion would receive support for up to two years. The total amount of the fixed cost is £329,345,366, and the total ongoing costs in the UK for the first 6 months are estimated to be £121,752,925, with an additional £13,684,204 for those children staying on a plan for at least 2 years.

**Table 3B:** Child social care costs due to maltreatment in 2015.

| **Table 3B: Child social care costs due to maltreatment in 2015** |  |  |  |  |  |  |
| --- | --- | --- | --- | --- | --- | --- |
| **Child social care** | **Percentage newly looked after children receiving intervention (%)** | **Number** | **Unit cost (£)** | **Duration (days)** | **Cost per child (£, discounted)** | **Total cost (£, discounted)** |
| Foster care | 78 | 14,518 | 88<br>(per day) | 785 | 67,672 | 982,460,996 |
| Local authority care | 14 | 2,609 | 455<br>(per day) | 785 | 349,895 | 912,886,189 |
| Child Protection Plan |  |  |  |  |  |  |
| Fixed cost<sup>1</sup> | 100 | 61,894 | 5,321<br>(per child) |  | 5,321 | 329,345,366 |
| Ongoing support costs (6 months) | 100 | 61,894 | 328<br>(per month) | First 6 months | 1,967 | 121,752,925 |
| Ongoing support costs after first 6 months | 3.7% England, Wales;<br>3.4% Scotland;<br>8.6% Northern Ireland | 2,372 | 328<br>(per month) | From month 7 to month 24 | 5,768 | 13,684,204 |

<sup>1</sup> Initial contact and referral, assessment, cost close case, core assessment, planning and review section 47 and public.

We assume that 78% of newly looked after children are put into foster care and 14% are placed in a local authority care home (including residential care homes, secure units, homes and hostels, and residential schools), based on English data for all newly-looked after children, irrespective of the cause in 2014 (Department of Education, 2015).^12^ We applied this to figures on the total number of newly looked after children in the UK that year. We assume that each child spends an average period of 785 days in foster care or local authority care, based on data from the same source. The costs of being placed for adoption, placement with parents, other placements in the community, and other placements are not known and therefore not included. We calculated the discounted cost per child receiving foster care, local authority care and multiplied these by the estimated numbers of newly looked after maltreated children receiving each intervention. The total discounted costs were £982,460,996 and £912,886,189 respectively (Table 3B).

The total cost of child social care is estimated to be £2,360,129,680. This figure was divided by the estimate of the incidence of child maltreatment in 2015 (61,894 (Department of Education, 2016))to give an estimate of the average cost per victim of £38,132.

#### Special education costs

There is evidence that maltreated children are more likely to receive special education support (Fang, Brown et al, 2012; Fisher, 2016). Fisher (2016) calculated that the proportion of children using special education services who were reported to have been severely maltreated (physical abuse, sexual abuse, emotional abuse or neglect or physical neglect) up to 12 years of age was 42%, compared to 20% among children who were not reported to be maltreated. Therefore, the incremental effect of child maltreatment on receipt of special education was 22 percentage points (unadjusted).

The Department for Children, Schools and Families reports that £3,310 per pupil was spent on the provision of education for children with special educational needs in England in 2008-2009 (DCSF January Schools Census 2009). Inflating this value to 2015 we assume every child incurred costs of £3,740 in 2015 for special education services.

We assume that provision of special education services starts at the age maltreatment starts and these costs are incurred until students are able to leave full-time education (assumed to be age 16 years). Hence, the duration of special education provision is assumed to be 10 years. The undiscounted cost per maltreated child in receipt of special education support is £32,195, and the cost per victim is estimated to be 0.22 * £32,195 = £7,068.

#### Productivity losses due to reduced employment

As discussed, in this category we include the costs of lost productivity due to reduced employment as a result of child maltreatment. Our NCDS analysis using the global CM measure showed that child maltreatment was negatively correlated with being in employment at ages 42 and 50 years, but the effects at 33 and 55 years were non-significant. This was also the case at age 23 (Table A5). We therefore assumed the marginal effect of child maltreatment on employment was zero at all ages up to 33 years and also from age 55 onwards. The effect of child maltreatment on the probability of being employed over the lifetime was calculated by assuming a straight line relation between marginal effects at ages 33, 42, 50 and 55 years.

Lost productivity due to reduced employment was valued using age-specific earnings before deduction of taxes in 2013-14 (the most recent year available) (ONS, 2014)^13^, inflated to 2015 prices. Based on previous studies (McCrone 2018) we assumed that earnings would increase by 2% per year. The NCDS analysis showed that child maltreatment did not have a significant effect on earnings so we did not account for this in our analysis.

We calculated the value of lifetime productivity losses due to reduced employment as the product of earnings at each age and the marginal effect of child maltreatment on employment. Future costs were discounted and summed across the lifetime. On this basis the average discounted lifetime productivity loss due per victim of child maltreatment was estimated to be £14,037.

#### Total costs and sensitivity analyses

We calculated our central estimate of the mean total lifetime costs of non-fatal child maltreatment per victim by summing the costs per victim across the various cost components described above. We also computed the proportion of the total accounted for by each component.

As indicated, there is uncertainty in our analysis; we therefore undertook several sensitivity analyses. First, we undertook a probabilistic sensitivity analysis (PSA) and used this to calculate 95% uncertainty intervals around the central estimate and the probability that the mean total lifetime costs of non-fatal child maltreatment per victim would be higher than a range of pre-specified values. The parameters we varied in the analysis were those we were able to assign probability distributions to, to reflect the uncertainty with each parameter value (Briggs, 2006) (Table A9). We used beta distributions to model uncertainty in the probabilities, and gamma and triangular distributions to model uncertainty in costs (gamma distributions were used where standard errors around mean values were known, triangular distributions were used where they were not; where minima and maxima were not known to parameterise triangular distributions we assumed values ±50% of the central estimate). A random value from the corresponding distribution for each parameter was selected. For each simulation we generated an estimate of the total lifetime costs of non-fatal child maltreatment per victim and each individual cost component. This was repeated 1000 times and the results for each simulation were noted. 95% uncertainty intervals were calculated as the 2.5^th^ and 97.5^th^ percentiles of the simulated values. The distribution of values was summarised graphically and the probability the total cost was greater a range of pre-specified values was calculated as the proportion of the simulations greater than values £0 to £200,000.

We also undertook univariate deterministic sensitivity analysis varying key values within plausible ranges one at a time. This was undertaken to highlight the sensitivity of the central estimates to individual parameter values. We varied the discount rate between 0% and 5% per annum. For unplanned injury-related admissions we assumed one unplanned admission per victim and that 31.2% would have a readmission (Wiljaars et al, 2016), and that each admission would be associated with it 11.5 outpatient visits based on the ratio of outpatient visits to inpatient admissions in the NHS,^14^ and an equivalent number of GP visits. For short-term mental health problems we varied the annual health care costs and criminal justice system costs per victim, and the impact of child maltreatment on these problems; we also examined the effect of using the short-term health-related cost estimates from Fang et al (2012) in the USA instead of our estimates. For long-term health-related costs we explored the impact of increasing the marginal effect of child maltreatment on each outcome, of extrapolating the effects to older and younger ages, and of using the estimates from the USA from Fang et al. (2012). For the criminal justice system costs we increased the number of cases ending in court proceedings and convictions, and used the estimates from the USA by Fang et al. (2012). For social care costs we increased the unit costs of child protection plans and monthly ongoing support, and used the estimates for child welfare costs from the USA by Fang et al. (2012). For special education costs we varied the unit cost of support and the marginal effect of child maltreatment. For reduced employment we varied the wages that would be earned to below the national average level, we varied the annual increase in wages, we explored the impact of increasing the (negative) marginal effect of child maltreatment on employment, of extrapolating the effects to older and younger ages, and of using the estimates from the USA from Fang et al. (2012). We judged that changes £10,000 higher or lower than the central estimate were substantive.

#### Lifetime costs per death from child maltreatment

Following Fang et al (Fang, Brown et al, 2012) we included two components to calculate the lifetime cost per victim of fatal child maltreatment: health care costs associated with fatal injuries; and, lifetime costs of lost productivity. We also included criminal justice system costs for perpetrators of fatal child maltreatment in sensitivity analyses. We have not been able to include costs associated with protecting other children in the family or in regular contact with perpetrators.

To calculate health care costs we used data from two published studies that calculated the cost of fatal blunt trauma and penetrating trauma injuries in the UK; mean costs per fatality for these injuries were £10,966 (Christensen, Ridley et al 2008) and £4,278, (Christensen, Nielsen et al 2008) respectively, in 2004/5 prices. These figures included the costs of transport to the hospital, hospital stays in the Accident and Emergency department, critical care unit and on the general ward, and the costs of procedures while in hospital. Updated to 2014/15 prices the values were £13,863 and £5,408, respectively. We used the lower of these two values in our central estimate and included the higher value in sensitivity analysis. To calculate lost productivity costs we used the human capital approach and multiplied figures for mean annual earnings by age (ONS, 2014) with employment rates by age (from age 16 to 67; (ONS, 2016)). The earnings figures were discounted to present value terms and inflated assuming a constant annual increase in earnings of 2% (McCrone 2008). The employment-adjusted earnings figures were summed across the lifetime to provide an estimate of total lifetime earnings accounting for the likelihood of employment. We assume this figure represents mean lifetime productivity and also the lost productivity that would be incurred with a fatality from maltreatment in childhood. This assumes lifetime earnings and employment for someone who died as a result of being maltreated as a child would equal national average figures if they had not died.

We did not include a probabilistic sensitivity analysis for the lifetime costs per death from child maltreatment (including 95% confidence intervals) as it was not possible to assign probability distributions to the key parameters. We undertook univariate deterministic sensitivity analysis we varied the discount rate, health care costs, and assumptions concerning the lost productivity. We also included criminal justice costs assuming all cases would result in court proceedings and a conviction.

---

**Table A1:** Definition of Child Maltreatment Used in the UK Literature.

| Study | Author | Data | Definition of maltreatment |
| --- | --- | --- | --- |
| Child Neglect and Maltreatment and Childhood-to-Adulthood Cognition and Mental Health in a Prospective Birth Cohort. | Geoffroy MC | NCDS | Neglect at 7 and 11 (parents and teacher): mother/father hardly ever takes on outings, had little interest in the child's education, child looks undernourished, scruffy/dirty. Neglect at age 45 (related to 16 years): I was neglected, Mother/father not at all affectionate toward me. Abuse at age 45 (related to 16 years) covers psychological abuse, physical abuse, sexual abuse and witnessing physical/sexual abuse. |
| Child maltreatment and household dysfunction: associations with pubertal development in a British birth cohort. | Li L | NCDS | Neglect at 7: mother/father hardly ever reads to child or takes on outings, had little interest in the child's education, low parental aspiration. Abuse at age 45 (physical, sexual, psychological), reported at 45 (related to 16). Witnessing abuse, reported at 45 (related to 16). Neglect at age 45 (related to 16): I was neglected, Mother/father not at all affectionate toward me. |
| The influence of childhood adversity on social relations and mental health at mid-life. | Ford E | NCDS | Neglected appearance at 7 and/or 11: the medical examiner reported an underfed/neglected. Parental physical abuse and sexual abuse reported at 45 years. Cumulative adversity was measured by counting the reports of neglected appearance, parental physical abuse, parental sexual abuse, but also being in care, parental divorce, maternal and paternal absence. |
| Does the influence of childhood adversity on psychopathology persist across the lifecourse? A 45-year prospective epidemiologic study. | Clark C | NCDS | Neglected appearance at 7 and/or 11: the medical examiner reported an underfed/neglected. Parental physical abuse and sexual abuse reported at 45 years. Cumulative adversity was measured by counting the reports of neglected appearance, parental physical abuse, parental sexual abuse, but also being in care, parental divorce, maternal and paternal absence. |
| Obesity and type 2 diabetes risk in midadult life: the role of childhood adversity. | Thomas C | NCDS | Neglect: little interest in child's education rated for each parent by teachers at 7, 11, 16; hardly ever has outings with mother or father at 7, 11; mother or father hardly ever reads to the child at 7; low parental aspirations at 11 and 16. Neglect at 7 and 11 (by teacher) "appearance of child" as scruffy or dirty. Emotional neglect at 45 (related to 16): how affectionate was mother/father. Physical neglect at 45 (related to 16): I was neglected. Abuse at 45 (related to 16): emotional, physical and sexual together. Witnessing abuse at 45 (related to 16). Received too much physical punishment defined as household dysfunction at 45 (related to 16) |
| Childhood maltreatment and BMI trajectories to mid-adult life: follow-up to age 50 y in a British birth cohort. | Power C | NCDS | Neglect at 7 and 11 (parents and teacher): mother/father hardly ever takes on outings, had little interest in the child's education, child looks undernourished, scruffy/dirty. Abuse at age 45 (related to 16 years) covers psychological abuse, physical abuse and sexual abuse. |
| Childhood adversity as a risk for cancer: findings from the 1958 British birth cohort study. | Kelly-Irving M | NCDS | Physical neglect at 7, 11, 16y: child appears undernourished/dirty at 7, 11, 16y. 'Adverse childhood experiences', however also include: being in care, offenders, parental separation, mental illness, alcohol abuse. |
| Adverse childhood experiences and physiological wear-and-tear in midlife: Findings from the 1958 BBC. | Barboza Soli-s C | NCDS | Physical neglect at 7, 11, 16y: child appears undernourished/dirty at 7, 11, 16y. 'Adverse childhood experiences', however also include: being in care, offenders, parental separation, mental illness, alcohol abuse. |
| Adverse childhood experiences and child-to-adult height trajectories in the 1958 British birth cohort. | Denholm R | NCDS | Neglect at 7: mother/father hardly ever reads to child or takes on outings, had little interest in the child's education, low parental aspiration. Neglect at age 45 (related to 16): I was neglected, Mother/father not at all affectionate toward me. Physical abuse, sexual abuse, psychological abuse, reported at 45 (related to 16). Witnessing abuse, reported at 45 (related to 16). |
| Child maltreatment and household dysfunction in a British birth cohort | Denholm R | NCDS | Psychological (or emotional) abuse: Intentional behaviour that conveys to a child that he/she is worthless, flawed, unloved, unwanted, endangered or valued only in meeting another's needs. *UK definition includes harmful (unintentional) parent-child interactions: 'the persistent emotional ill-treatment of a child such as to cause severe and persistent adverse effects on the child's emotional development'*. Verbally abused at age 45 (related to 16): I was verbally abused by a parent (45 years); I suffered humiliation, ridicule, bullying or mental cruelty from a parent. Physical abuse: Intentional use of physical force or implements against a child that results in, or has the potential to result in, physical injury. Physical abuse at 45 (related to 16): I was physically abused by a parent – punched, kicked or hit or beaten with an object, or needed medical treatment. Sexual abuse: Any completed or attempted sexual act, sexual contact or non-contact sexual interaction with a child by a caregiver. Sexually abused at 45 (related to 16): I was sexually abused by a parent. Witnessing intimate partner violence: any incident of threatening behaviour, violence or abuse (psychological, physical, sexual, financial or emotional) between intimate partners or adult family members, irrespective of sex or sexuality. Witness of violence at 45 (related to 16): I witnessed physical or sexual abuse of others in my family. Neglect: Failure to meet a child's basic physical, emotional, medical/dental or education need; failure to provide adequate nutrition, hygiene or shelter; or failure to ensure a child's safety. Neglected at age 45 (related to 16): I was neglected; Mother unaffectionate; Father unaffectionate; Mother hardly ever reads to child (7 years); Father hardly ever reads to child (7 years). |
| Childhood abuse is associated with methylation of multiple loci in adult DNA. | Suderman M | NCDS | Abuse reported at 45 (related to 16). Emotional: I was verbally abused by a parent; I suffered humiliation, ridicule, bullying or mental cruelty from a parent. Physical: I was physically abused by a parent – punched, kicked or hit or beaten with an object, or needed medical treatment. Sexual: I was sexually abused by a parent. |
| Understanding the mechanisms through which adverse childhood experiences affect lifetime economic outcomes. | Schurer S | NCDS | Adverse Childhood Experiences Index comprised of 6 different items, including physical neglect "whether the child appears undernourished or dirty at ages 7 or 11", from teacher responses to the Bristol Social Adjustment Guide. |
| Maternal childhood abuse and offspring adjustment over time. | Collishaw S | ALSPAC | Maternal childhood experience of abuse (sexual, physical, and emotional), retrospectively: "whether a parent had been physically or emotionally cruel to them prior to age 17 years". |
| Child maltreatment in the children of the nineties": a cohort study of risk factors." | Sidebotham P | ALSPAC | Social Services child protection registers were screened for any children with birth dates who have been investigated for possible child abuse or neglect. Physical injury: actual or likely physical injury to a child, or failure to prevent physical injury (or suffering) to a child including deliberate poisoning, suffocation and Munchausen syndrome by proxy. Neglect: persistent/severe neglect of a child, or failure to protect a child from exposure to any kind of danger, including cold or starvation, or extreme failure to carry out important aspects of care, resulting in significant impairment of the child's health or development, including non-organic failure to thrive. Sexual abuse: actual or likely sexual exploitation of a child or adolescent. The child may be dependent and/or developmentally immature. Emotional abuse: actual or likely severe adverse effect on the emotional and behavioural development of a child caused by persistent or severe emotional ill-treatment or rejection. All abuse involves some emotional ill-treatment. |
| Child maltreatment in the children of the 90s: the child. | Sidebotham P. | ALSPAC | Social Services child protection registers were screened for any children who have been investigated for possible child abuse or neglect. |
| Child maltreatment in the Children of the 90s: deprivation. | Sidebotham P. | ALSPAC | Social Services child protection registers were screened for any children who have been investigated for possible child abuse or neglect. |
| Child maltreatment in the children of the 90s: a longitudinal study of parental risk factors. | Sidebotham P. | ALSPAC | Social Services child protection registers were screened for any children who have been investigated for possible child abuse or neglect. |
| Measuring adolescents' exposure to victimization: the Environmental Risk (E-Risk) Longitudinal Twin Study | Fisher, Helen L. | E-Risk | Sexual victimization, family violence, maltreatment, neglect. |
| Childhood Trauma and Children's Emerging Psychotic Symptoms: A Genetically Sensitive Longitudinal Cohort Study | Arseneault L | E-Risk | Maltreatment by an adult by interviewing mothers. Do you remember when the child was disciplined severely enough that s/he may have been hurt? Did you worry that you or someone else may have harmed or hurt the child during those years? |
| The limits of child effects: evidence for genetically mediated child effects on corporal punishment but not on physical maltreatment. | Jaffee SR | E-Risk | Corporal punishment and child physical maltreatment. Whether the child had experienced a variety of disciplinary practices in the past year, some of which assessed nonphysical discipline (e.g., "isolation," "withdrew privileges") and three of which assessed corporal punishment ("grabbing or shaking," "smacking or hitting," or engaging in "other physical discipline"). Probe questions about potential physical maltreatment, including "When [name] was a toddler, do you remember any time when she/he was disciplined severely enough that she/he may have been hurt?" and "Did you worry that you or someone else [such as a babysitter, a relative, or a neighbour] may have harmed or hurt [name] during those years?" |
| Variation in recorded child maltreatment concerns in UK primary care records: a cohort study using The Health Improvement Network (THIN) database. | Woodman J | THIN | "Maltreatment-related" codes. 1. Child protection procedures. Codes indicating child protection plan, case conference, or child protection investigation. 2. Direct references to maltreatment or out-of-home-care. Codes making explicit reference to abuse or neglect (including domestic violence) or to formal out-of-home care. 3. High risk child. Codes indicating high levels of social welfare need or concern in the child or family, including a history of abuse or neglect. 4. Contact with children's social care. Codes indicating that the child is involved with or has been referred to children's social care (not including codes specifically referring to 'child protection procedure' codes. |

**Table A2:** Marginal effect of child maltreatment on the probability of mental health problems.

| **Disorder** | **Children age 5-10 years** | **Children age 11-15 years** |
| --- | --- | --- |
| Emotional disorder | 0.080 | 0.059 |
| Conduct disorder | 0.315 | 0.345 |
| Hyperkinetic disorder | 0.091 | 0.061 |

**Table A3:** Child social care costs due to maltreatment in 2015.

| **Child social care** | **Percentage newly looked after children receiving intervention (%)** | **Exact Number** | **Unit cost (£)** | **Duration (days)** | **Cost per child (£, discounted)** | **Total cost (£, discounted)** |
| --- | --- | --- | --- | --- | --- | --- |
| Foster care | 78 | 14,518 | 88<br>(per day) | 785 | 67,672 | 982,460,996 |
| Local authority care | 14 | 2,609 | 455<br>(per day) | 785 | 349,895 | 912,886,189 |
| **Child Protection Plan** |  |  |  |  |  |  |
| Fixed cost<sup>1</sup> | 100 | 61,894 | 5,321<br>(per child) |  | 5,321 | 329,345,366 |
| Ongoing support costs (6 months) | 100 | 61,894 | 328<br>(per month) | First 6 months | 1,967 | 121,752,925 |
| Ongoing support costs after first 6 months | 3.7% England, Wales;<br>3.4% Scotland;<br>8.6% Northern Ireland | 2,372 | 328<br>(per month) | From month 7 to month 24 | 5,768 | 13,684,204 |

<sup>1</sup>Initial contact and referral, assessment, cost to close case, core assessment, planning and review section 47 and public law outline.

**Table A4:**
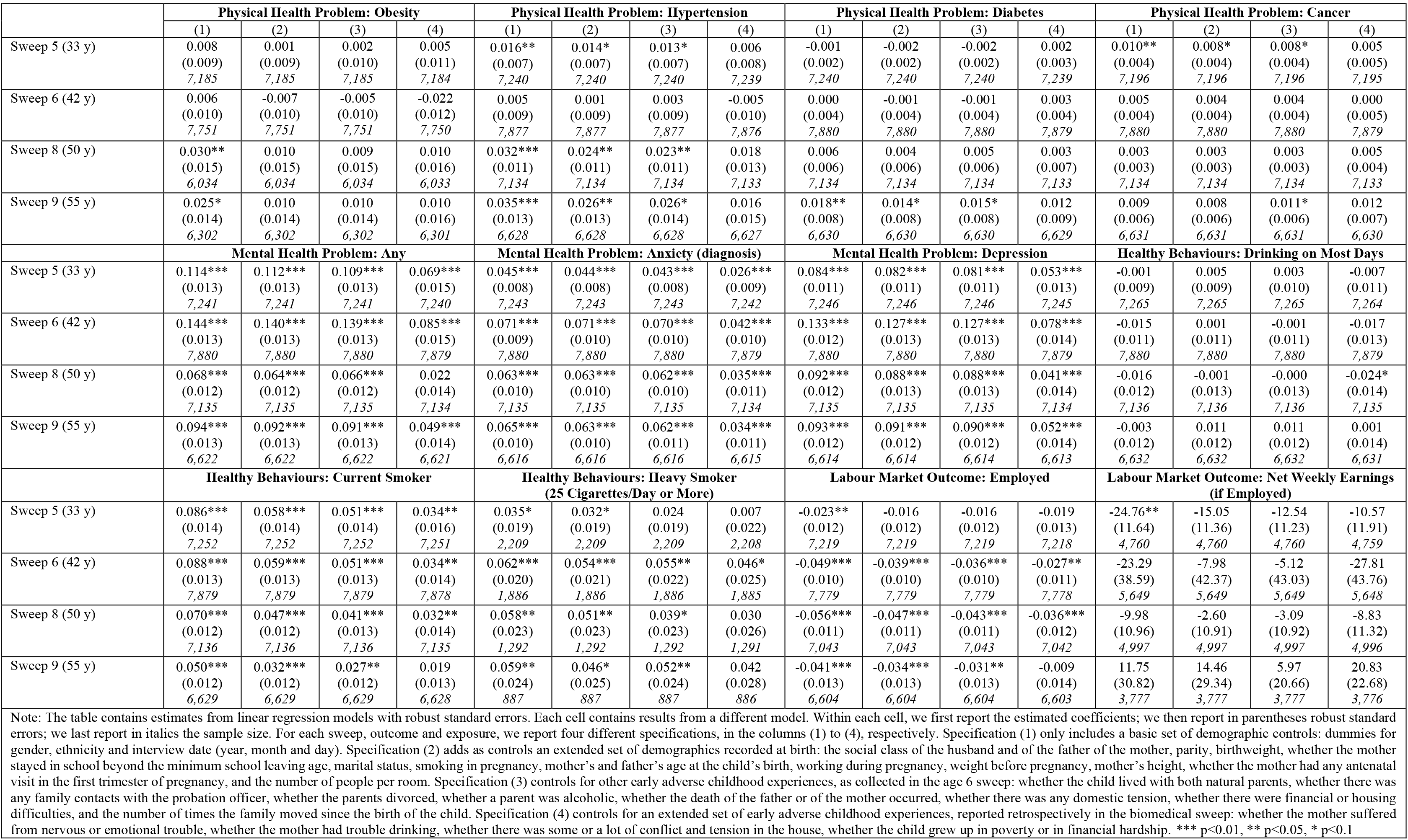
NCDS Results of the Effects of the global measure of CM.

**Table A5:** Supplementary NCDS Results of the Effects of the Global Measure of CM.

|  | Labour Market Outcome:<br>Employed | Healthy Behaviours:<br>Current Smoker | Mental Health Problem:<br>Any | Mental Health Problem:<br>Neurotic disorder<sup>1</sup> | Mental Health Problem:<br>Depression |
| --- | --- | --- | --- | --- | --- |
| Sweep 4 (Age 23) | -0.0202<br>(0.0147)<br>*7,105* | 0.0229<br>(0.0201)<br>*4,865* | 0.0059<br>(0.0051)<br>*7,105* | 0.0085*<br>(0.0048)<br>*7,105* | -0.0010<br>(0.0053)<br>*7,105* |

**Table A6:** ELSA Results of the Effects of the global measure of CM.

|  | (1) | (2) |
| --- | --- | --- |
|  | **Physical Health Problem: Obesity** |  |
| All ages | 0.019 (0.014) | 0.023 (0.015) |
|  | **Physical Health Problem: Hypertension** |  |
| All ages | 0.015 (0.013) | 0.017 (0.014) |
|  | **Physical Health Problem: Diabetes** |  |
| All ages | 0.009 (0.008) | 0.010 (0.009) |
|  | **Physical Health Problem: Cancer** |  |
| All ages | 0.001 (0.007) | 0.001 (0.007) |
|  | **Mental Health Problem: Emotional/psychotic disorder (all types)** |  |
| All ages | 0.069*** (0.009) | 0.045*** (0.010) |
| By age 30 | 0.013** (0.005) | 0.008 (0.005) |
| By age 40 | 0.022*** (0.006) | 0.014** (0.006) |
| By age 50 | 0.042*** (0.008) | 0.026*** (0.008) |
| By age 60 | 0.050*** (0.009) | 0.033*** (0.009) |
| By age 70 | 0.051*** (0.009) | 0.034*** (0.009) |
|  | **Mental Health Problem: Anxiety** |  |
| All ages | 0.056*** (0.008) | 0.037*** (0.008) |
| By age 30 | 0.010** (0.004) | 0.007 (0.005) |
| By age 40 | 0.018*** (0.005) | 0.012** (0.006) |
| By age 50 | 0.032*** (0.007) | 0.020*** (0.007) |
| By age 60 | 0.038*** (0.007) | 0.025*** (0.008) |
| By age 70 | 0.039*** (0.007) | 0.026*** (0.008) |
|  | **Mental Health Problem: Depression** |  |
| All ages | 0.058*** (0.009) | 0.038*** (0.009) |
| By age 30 | 0.013*** (0.005) | 0.009* (0.005) |
| By age 40 | 0.022*** (0.006) | 0.015** (0.006) |
| By age 50 | 0.039*** (0.007) | 0.022*** (0.007) |
| By age 60 | 0.046*** (0.008) | 0.030*** (0.008) |
| By age 70 | 0.047*** (0.008) | 0.031*** (0.008) |

Note: The table contains estimates from linear regression models (robust standard errors in parentheses). We report results for two different specifications. Specification (1) (left column) only includes as controls a basic set of demographics: age, gender, ethnicity and the month and year of the interview. Specification (2) (right column) adds a set of adverse early childhood experiences: self-reported health in childhood, whether missed school for over a month due to health as child, whether in hospital for more than a month due to health as child, whether when aged <16 parents ever permanently separated or divorced, whether when aged <16 either parents unemployed for over 6mths when wanted to work, whether when aged <16 parents argue or fight very often, whether when aged <16 ever separated from mother for 6 months or more, whether when aged <16 parents drunk/took drugs/had mental health problems.

**Table A7:** ELSA Results of the Effects of the global measure of CM.

|  | (1) | (2) |
| --- | --- | --- |
|  | **Healthy Behaviours: Heavy Drinking** |  |
| All ages | 0.026*** (0.009) | 0.029*** (0.009) |
|  | **Healthy Behaviours: Current Smoker** |  |
| All ages | 0.016* (0.009) | 0.006 (0.010) |
|  | **Healthy Behaviours: Heavy Smoker (25 Cigarettes/Day or More)** |  |
| All ages | 0.012 (0.015) | 0.010 (0.016) |
|  | **Labour Market Outcome: Employed** |  |
| All ages | -0.017* (0.009) | -0.010 (0.010) |
|  | **Labour Market Outcome: Weekly Earnings (if Employed)** |  |
| All ages | -8.17 (10.02) | 1.95 (9.94) |
|  | **Labour Market Outcome: Weekly Earnings (All)** |  |
| All ages | -18.73 (12.46) | -6.98 (12.94) |
|  | **Labour Market Outcome: Weekly Disability-Related Benefits (if Participants)** |  |
| All ages | 12.47 (9.31) | 14.02 (13.79) |
|  | **Labour Market Outcome: Weekly Disability-Related Benefits (All)** |  |
| All ages | 47.49*** (16.18) | 35.59** (15.60) |

Note: The table contains estimates from linear regression models with robust standard errors, apart from the outcomes “weekly earnings (all)” and “weekly disability-related benefits (all)”, which are estimated using tobit models. Each cell contains results from a different model. We report two different specifications, in the columns (1) to (2), respectively. Specification (1) only includes as controls a basic set of demographics: age, gender, ethnicity and the month and year of the interview. Specification (2) adds a set of adverse early childhood experiences: self-reported health in childhood, whether missed school for over a month due to health as child, whether in hospital for more than a month due to health as child, whether when aged <16 parents ever permanently separated or divorced, whether when aged <16 either parents unemployed for over 6mths when wanted to work, whether when aged <16 parents argue or fight very often, whether when aged <16 ever separated from mother for 6 months or more, whether when aged <16 parents drunk/took drugs/had mental health problems.

**Table A8:** Exact Definitions of the Outcome Variables in the NCDS.

| **Table A8: Exact Definitions of the Outcome Variables in the NCDS** |  |  |  |  |
| --- | --- | --- | --- | --- |
| Outcome | Sweep 5 | Sweep 6 | Sweep 8 | Sweep 9 |
| **Physical health problems** |  |  |  |  |
| Obesity | BMI=>30 | BMI=>30 | BMI=>30 | BMI=>30 |
| Diabetes<br>(diagnosis,<br>general) | Ever suffered diabetes | Ever had diabetes | Suffers diabetes | Health problems: diabetes |
| Hypertension<br>(diagnosis) | Ever suffered high blood pressure | Ever had high blood pressure | Suffers high blood pressure | Health problems: high blood pressure |
| Cancer<br>(diagnosis, all types) | Ever suffered cancer | Ever had cancer | Suffers cancer or leukaemia | Health problems: cancer or leukaemia |
| **Mental health problems** |  |  |  |  |
| Mental problems | Ever suffered emotional problem and seen a specialist | Seen specialist for any mental disorder | Seen a doctor for any mental disorder | Health problems: depression, emotional and psychiatric |
| Anxiety<br>(diagnosis) | Ever feel jittery/anxious and seen a specialist | Seen specialist for feeling jittery/anxious | Seen a doctor for feeling jittery/anxious | Health problems: feeling jittery/anxious |
| Depression<br>(diagnosis) | Ever feel low/depressed/sad and seen a specialist | Seen specialist for feeling low/depressed/sad | Seen a doctor for feeling low/depressed/sad | Health problems: feeling low/depressed/sad |

**Table A9:** Variables, distributions and parameters included in probabilistic sensitivity analysis.

| | Central value | SE | Distribution | Lower limit | Upper limit | $\alpha$ | $\beta$ |
| --- | --- | --- | --- | --- | --- | --- | --- |
| Short-term health-related costs |  |  |  |  |  |  |  |
| Unplanned hospital admissions for injuries |  |  |  |  |  |  |  |
| MVR rate <1 year | 102.8 | 3.878 | Gamma |  |  | 702.84 | 0.15 |
| MVR rate 1-10 years | 23.3 | 0.612 | Gamma |  |  | 1448.26 | 0.02 |
| MVR rate 11-18 years | 103.3 | 1.378 | Gamma |  |  | 5623.02 | 0.02 |
| Unit cost per admission | £853 |  | Triangular | 560 | 1014 |  |  |
| Short-term mental health problems |  |  |  |  |  |  |  |
| Mean annual cost of disorders in children |  |  |  |  |  |  |  |
| Hyperkinetic disorder | 3,605 | 783 | Gamma |  |  | 21.15 | 170.48 |
| Conduct disorder | 2,153 | 526 | Gamma |  |  | 16.75 | 128.52 |
| Emotional disorder | 1,352 | 177 | Gamma |  |  | 58.21 | 23.22 |
| Annual criminal justice cost for children with conduct disorder | 3,436 |  | Triangular | 1718 | 5153 |  |  |
| Marginal effect of child maltreatment on probability of mental health disorders in children aged 5-10 |  |  |  |  |  |  |  |
| Emotional disorder | 0.08 |  | Beta | 0 | 1 | 38.28 | 309.72 |
| Conduct disorder | 0.315 |  | Beta | 0 | 1 | 127.02 | 220.98 |
| Hyperkinetic disorder | 0.091 |  | Beta | 0 | 1 | 38.628 | 309.372 |
| Marginal effect of child maltreatment on probability of mental health disorders in children aged 11-15 |  |  |  |  |  |  |  |
| Emotional disorder | 0.059 |  | Beta | 0 | 1 | 57.12 | 422.88 |
| Conduct disorder | 0.345 |  | Beta | 0 | 1 | 194.4 | 285.6 |
| Hyperkinetic disorder | 0.061 |  | Beta | 0 | 1 | 34.08 | 445.92 |
| Criminal justice costs incurred by perpetrators |  |  |  |  |  |  |  |
| Unit cost of court proceedings | 11,746 |  | Triangular | 5873 | 17620 |  |  |
| Unit cost of convictions | 20,362 |  | Triangular | 10181 | 30543 |  |  |
| Unit cost for sex offenders treatment programmes | 11,025 |  | Triangular | 5513 | 16538 |  |  |
| Proportion of cases ending in court proceedings | 0.176 |  | Beta | 0 | 1 | 4100 | 19200 |
| Proportion of cases ending in convictions | 0.107 |  | Beta | 0 | 1 | 2500 | 20800 |
| Proportion of cases of sexual abuse ending in sex offenders treatment programmes | 0.048 |  | Beta | 0 | 1 | 1118 | 22182 |
| Social care costs |  |  |  |  |  |  |  |
| Foster care cost per day | 88 |  | Triangular | 44 | 132 |  |  |
| Local Authority care cost per day | 455 |  | Triangular | 228 | 683 |  |  |
| Unit cost of Child Protection Plan | 5,321 |  | Triangular | 2661 | 7982 |  |  |
| Monthly child care support cost | 328 |  | Triangular | 164 | 492 |  |  |
| Proportion newly looked after children receiving intervention in foster care | 0.777 |  | Beta | 0 | 1 | 24150 | 6920 |
| Proportion newly looked after children receiving intervention in Local Authority care | 0.139 |  | Beta | 0 | 1 | 4340 | 26730 |
| Special education costs |  |  |  |  |  |  |  |
| Unit cost of special educational support per victim | 3,740 |  | Triangular | 1870 | 5610 |  |  |
| Marginal effect of child maltreatment on receipt of special education support | 0.22 |  | Beta | 0 | 1 | 34 | 47 |
| Long-term health-related costs |  |  |  |  |  |  |  |
| Annual cost of disorders |  |  |  |  |  |  |  |
| Depression (NHS costs only) | 2,417 |  | Triangular | 1209 | 3627 |  |  |
| Depression (productivity costs) | 4,478 |  | Triangular | 2239 | 6718 |  |  |
| Anxiety (NHS costs only) | 1,282 |  | Triangular | 641 | 1923 |  |  |
| Anxiety (productivity costs) | 662 |  | Triangular | 331 | 992 |  |  |
| Smoking (NHS and productivity costs) | 1,805 |  | Triangular | 902 | 2707 |  |  |
| Smoking (NHS costs only) | 430 |  | Triangular | 215 | 645 |  |  |
| Alcohol abuse (NHS costs only) | 211 |  | Triangular | 106 | 317 |  |  |
| Alcohol abuse (criminal justice costs) | 1,822 |  | Triangular | 911 | 2734 |  |  |
| Alcohol abuse (productivity costs) | 529 |  | Triangular | 265 | 794 |  |  |
| Marginal effect of child maltreatment on probability of disorders |  |  |  |  |  |  |  |
| Depression at age 33 | 0.053 | 0.013 | Gamma |  |  | 17.14 | 0.00 |
| Depression at age 42 | 0.078 | 0.014 | Gamma |  |  | 29.19 | 0.00 |
| Depression at age 50 | 0.041 | 0.014 | Gamma |  |  | 8.27 | 0.00 |
| Depression at age 55 | 0.052 | 0.014 | Gamma |  |  | 14.20 | 0.00 |
| Anxiety |  |  |  |  |  |  |  |
| Anxiety at age 33 | 0.026 | 0.009 | Gamma |  |  | 8.63 | 0.00 |
| Anxiety at age 42 | 0.042 | 0.010 | Gamma |  |  | 16.54 | 0.00 |
| Anxiety at age 50 | 0.035 | 0.011 | Gamma |  |  | 10.14 | 0.00 |
| Anxiety at age 55 | 0.034 | 0.011 | Gamma |  |  | 9.06 | 0.00 |
| Smoking |  |  |  |  |  |  |  |
| Current smoking at age 33 | 0.034 | 0.016 | Gamma |  |  | 4.64 | 0.01 |
| Current smoking at age 42 | 0.034 | 0.014 | Gamma |  |  | 5.60 | 0.01 |
| Current smoking at age 50 | 0.032 | 0.014 | Gamma |  |  | 5.45 | 0.01 |
| Alcohol |  |  |  |  |  |  |  |
| Heavy drinking at age 50 or more | 0.029 | 0.009 | Gamma |  |  | 9.57 | 0.00 |
| Reduced employment |  |  |  |  |  |  |  |
| Marginal effect of child maltreatment on probability of being employed |  |  |  |  |  |  |  |
| Employed at age 42 | -0.027 | 0.011 | Gamma |  |  | 5.67 | 0.00 |
| Employed at age 50 | -0.036 | 0.012 | Gamma |  |  | 8.42 | 0.00 |

All costs are in 2015 UK£. SE = standard error. MVR = maltreatment or violence-related.

## Footnotes

2 https://explore-education-statistics.service.gov.uk/find-statistics/serious-incident-notifications

3 Bald et al. (2019) and Gross and Baron (2021) use the same instrument and find that removal, instead, significantly increases test scores, improves children’s safety and educational outcomes and reduces grade repetition for girls.

4 Note that the datasets used in more recent papers, as noted in the first literature review, were not available to us when we started this project.

5 This reflects the age children become subject to a child protection plan or come on to the child protection register in the UK, and it is likely an upper bound; we did not find any reliable data on the age at which maltreatment starts.

6 Private communication from H. Fisher, based on published data from the E-Risk Twins.

7 The precise definitions of the outcome variables are reported in Table A8 of the Supplementary Material.

8 For example, the 119 notified child deaths (as part of the serious incident notifications reported to the Department for Education in England during the first half of 2020-21) will cost £940,758*119=£105,364,896.

1 A separate variable has been included as control in all the estimated models in case the answers include both “I had no mother/father figure” and “can’t say”.

2 We kept categories ‘No father/mother figure’ and ‘Can’t say’ separately from affirmative or negative answers to CM questions. In other words, for each CM measure, in the regression analyses the baseline category was not suffering CM, compared each of categories 1) ‘suffering CM’; 2) ‘No father/mother figure’; 3) ‘Can’t say’. In the paper we present results for (1); results for (2) and (3) are available upon request.

3 A separate variable has been included as control in all the estimated models in case the answers include both “no” and “can’t say”.

4 A separate variable has been included as control in all the estimated models in case the answer is “can’t say”.

5 A transfer payment (or government transfer) is a redistribution of income. These payments do not directly absorb resources or create output, and are made without any exchange of goods or services. Disability benefits are transfer payment as they represent a payment made or an income received in which no goods or services are being paid for. The primary reason for excluding transfer payments is to avoid “double counting” as these payments are made from the government and received from beneficiaries and therefor there is no loss from a societal perspective.

6 https://www.gov.uk/government/uploads/system/uploads/attachment_data/file/490866/GDP_Deflators_Qtrly_National_Accounts_December_2015_update.csv/preview

7 Department for Education (DfE) 2016

8 https://www.nice.org.uk/article/pmg9/chapter/5-The-reference-case#discounting

9 https://www.gov.uk/government/uploads/system/uploads/attachment_data/file/220541/green_book_complete.pdf

10 www.ash.org.uk/localtoolkit/docs/Reckoner.xls

11 http://www.thisismoney.co.uk/money/pensions/article-1679780/New-state-pension-age-retire.htm

12 https://www.gov.uk/government/uploads/system/uploads/attachment_data/file/464756/SFR34_2015_Text.pdf https://www.gov.uk/government/statistics/children-looked-after-in-england-including-adoption-2014-to-2015

13 https://www.gov.uk/government/statistics/distribution-of-median-and-mean-income-and-tax-by-age-range-and-gender-2010-to-2011

14 https://www.england.nhs.uk/statistics/statistical-work-areas/hospital-activity/monthly-hospital-activity/

